# Interventions to manage intolerance among patients prescribed statins for primary prevention of cardiovascular diseases: A systematic review and meta-analysis

**DOI:** 10.64898/2026.02.23.26346865

**Authors:** Shagoofa Rakhshanda, Joel Rhee, Siaw-Teng Liaw, Kerry-Anne Rye, Jitendra Jonnagaddala

## Abstract

The objective of this systematic review and meta-analysis was to identify the interventions used to manage intolerance in patients receiving statins for primary prevention of CVD and to determine the effectiveness of these interventions. This study was conducted according to the PRISMA checklist. The electronic databases MEDLINE (PubMed), SCOPUS, EMBASE, and CINAHL were searched for studies published until June 2025. Based on the NLA definition of statin intolerance, the outcomes were split into adverse effects caused by statins and statin discontinuation. In total, 1,238 studies were identified and screened. Nine studies were eligible for systematic review, and six studies were eligible for meta-analysis. The identified intervention strategies were adjuvant therapy, statin titration, replacing statins with other lipid-lowering agents and switching to different statin. The meta-analysis showed that the pooled risk ratio (RR) relative to control was 0.97 (95% CI, 0.86–1.08) in randomized controlled trials and 0.94 (95% CI, 0.63–1.42) in overall, with point estimates in favour of intervention arms. Moderate to substantial heterogeneity was observed, with I^2^ between 27% to 57%. Due to the smaller number of studies, no clear conclusions can be drawn regarding how the implemented interventions may affect statin discontinuation. This study showed no strong evidence that the implemented interventions reduced statin intolerance.

**PROSPERO registration number:** CRD42024587573

## 1. Introduction

Cardiovascular disease (CVD) is a condition that causes high rates of morbidity and mortality worldwide. In 2019, 32% of deaths worldwide were caused by CVD. Atherosclerotic cardiovascular disease (ASCVD) is a type of CVD that occurs due to the formation of cholesterol-rich plaques on the inner walls of arteries (intima), resulting in narrowing of the arterial lumen (1, 2, 3, 4). Statins are lipid-lowering drugs that lower cholesterol levels and are widely used as first-line pharmacological agents for the primary and secondary prevention of CVD, primarily ASCVD (5). Primary prevention measures seek to prevent the development of CVD before it occurs, whereas secondary prevention measures seek to prevent the future occurrence of CV events in patients with established disease (6). The effectiveness of statins as a preventive measure depends greatly on patients’ tolerance of statins, which in turn is associated with patients’ adherence to statin therapy (7).

Intolerance to statins is a clinical issue that is characterised by muscle symptoms and other adverse effects such as myopathy and rhabdomyolysis. Statin intolerance attenuates cholesterol lowering and increases cardiovascular risk (7). The estimated incidence rate of statin intolerance varies between 7% and 29%, leading to statin discontinuation (8). This variation in estimates is partly attributed to different definitions of intolerance including statin-associated muscle symptoms (SAMS) (9, 10). Other definitions include the inability to tolerate a statin dose, or two or more statins at any dose, limiting the effectiveness of the treatment (11, 12, 13). One commonly used definitions of statin intolerance, provided by the National Lipid Association (NLA), describes it as one or more adverse effects associated with statin therapy that improve with dose reduction or discontinuation (14, 15). However, in clinical practice, it is difficult to differentiate muscle symptoms caused by statin intolerance from those caused by other factors, such as vitamin D deficiency, hypothyroidism, and drug interactions, which can also cause muscle symptoms (16, 17).

Evidence suggests that there are several approaches for the management of statin intolerance, including changing dosages, switching to different statins, or substituting or adding non-statin lipid-lowering agents (18). However, owing to the complexity of statin intolerance in terms of adverse effects caused by statins (such as adverse events, muscle symptoms and myalgia) and statin discontinuation, there is a need for a comprehensive and deeper understanding of the management strategies. Such understanding may provide fundamental insights into statin adherence and CVD risk reduction. Therefore, the aim of this systematic review and meta-analysis was to identify and assess the effect of interventions used to manage statin intolerance in patients receiving statins for the primary prevention of CVD.

## 2. Methods

This systematic review and meta-analysis were conducted according to the Preferred Reporting Items for Systematic Reviews and Meta-Analyses (PRISMA) checklist **(Supplement 1)**. The protocol registration number for PROSPERO is CRD42024587573 (19). Previous publications reflect the initial findings based on this protocol (20, 21).

### 2.1 Eligibility criteria

This systematic review and meta-analysis included studies on patients prescribed statins for primary prevention of CVD. The eligibility criteria were developed using the Population, Intervention, Comparator, Outcome, and Study (PICOS) framework. Randomized controlled trials and non-randomized studies of interventions were eligible for inclusion. The included population comprised adults aged ≥ 18 years who were prescribed statins for primary prevention at a primary healthcare facility. The included studies evaluated interventions that managed statin intolerance among patients prescribed statins for primary prevention of CVD. The included interventions had to target patients receiving only statins or statins in combination with other medications for the primary prevention of CVD. The included comparators (controls) were placebo, statin monotherapy, or usual care groups (with no intervention or change in usual care activities). The outcomes of interest were measures of statin intolerance, including the number of patients with adverse events, adverse effects, serious adverse events, muscle-related symptoms, myalgia, and creatine kinase (CK) levels > 10 times the upper limit of normal (ULN). The included studies were clinical trials, intervention studies, and cohort studies investigating interventions that managed statin intolerance and were published in English language in peer-reviewed journals until June 2025. Studies not published in English language were excluded because of language limitations among the review team. Further details on the eligibility criteria are provided in **Supplement 2**.

### 2.2 Search strategy

The electronic databases MEDLINE (PubMed), EMBASE, SCOPUS, and CINAHL were searched for studies published until June 2025. The search terms are presented in **Supplement 3**.

### 2.3 Data collection

Based on the eligibility criteria, two researchers (SR and JJ) independently screened the titles and abstracts of all records to identify potentially relevant studies. The same two researchers (SR and JJ) independently screened the full texts screening of potentially relevant studies to determine eligibility. An automated software package (Covidence) was used for this purpose (22, 23).

The researchers (SR and JJ) independently extracted data from each eligible study using a data extraction sheet. The extracted data included details of the study, including the methods, patient characteristics, interventions, controls, and outcomes. Any discrepancies or differences in opinions were discussed and resolved during the study identification and data extraction processes. The interventions identified in this study were categorised based on therapeutic options for the management of statin intolerance, including switching to different statins, statin titration (dose alteration, frequency alteration, or intensity alteration), replacing statins with other lipid-lowering agents (such as ezetimibe, evolocumab, bempedoic acid, and proprotein convertase subtilisin/kexin type 9 [PCSK9] inhibitors), and adjuvant therapy (defined as the addition of non-statin drugs or lipid-lowering agents alongside statin therapy) (24, 25, 26).

### 2.4 Risk of bias assessment

We used the Cochrane Risk of Bias tool for randomized trials (RoB 2) to assess risk of bias in randomized controlled trials (RCTs) and the Cochrane risk of bias in non-randomized studies of interventions (ROBINS-I) to assess risk of bias in non-randomized studies of interventions (NRSI). Both RoB 2 and ROBINS-I included the following domains: bias due to deviations from intended interventions, bias due to missing outcome data, bias in measurement of outcome, and bias in selection of reported results. Additionally, RoB 2 included a domain for the bias arising from the randomization process, and ROBINS-I included domains for the bias due to confounding, bias in selection of participants into the study and bias in classification of interventions.

Two researchers (SR and JJ) independently assessed the studies and any discrepancies or differences in opinions were discussed and resolved. Publication bias was assessed using funnel plots, through which asymmetry was assessed visually (27). Egger’s test was not performed because fewer than 10 studies eligible for meta-analysis, consistent with Cochrane Handbook recommendations regarding tests for funnel plot asymmetry.

### 2.5 Data synthesis and analysis

Inconsistencies in the data were addressed through data synthesis, which reduced bias and increased generalisability, such as categorising all non-randomized trials, quasi-experimental studies and cohort studies as NRSIs. Data synthesis was guided by the intervention strategies that were used during the data collection phase. Meta-analysis was conducted when sufficient and relevant data were available. Data analysis using baseline and endline measurements was performed using the ‘meta’ package in R, with the overall effect size estimated using a random effects model (28, 29). Intolerance outcomes were measured using various approaches, such as adverse events, adverse effects, muscle-related symptoms, myalgia, and CK levels > 10 time the ULN. Variability in outcome measures were addressed by grouping comparable measures where appropriate. For example, the number of participants experiencing adverse effects caused by statins (such as adverse events, severe adverse events, muscle symptoms or myalgia) were categorised under the broader outcome of statin intolerance. Based on the NLA definition of statin intolerance, the outcomes were classified into two categories: adverse effects caused by statins or statin intolerance (as described above), and statin discontinuation (when patients discontinued statins because of intolerance). The estimated overall effect size was reported as the risk ratio (RR) with 95% confidence intervals (CIs). Cochran’s Q test, I^2^ statistic, and chi-square test were used to assess heterogeneity between studies. To explore whether effect estimates differed according to outcome definitions, subgroup analyses by intolerance measurement type were performed. Several other subgroup analyses were performed based on study designs and included analysis of the effectiveness of intervention strategies, as well as mean ages and locations of the participants. All analyses were stratified by study design. Tables and narrative reports were used to present the results of the systematic review and forest plots were used to present the results of the meta-analyses. A detailed analysis plan is available in a previously published protocol (19).

### 2.6 Ethics statement

This study is a systematic review and meta-analysis. It does not involve human participants.

## 3. Results

### 3.1 Selection of studies

The search of electronic databases yielded 1,238 studies, 190 of which were duplicates. Of the remaining 1,048 studies, 952 were excluded based on the eligibility criteria. This left 96 studies for full-text screening, of which 87 were excluded, and 9 were eligible for data extraction **(Figure 1)**. The full list of the 87 excluded studies is provided in **Supplement 4**.

**Figure 1:**
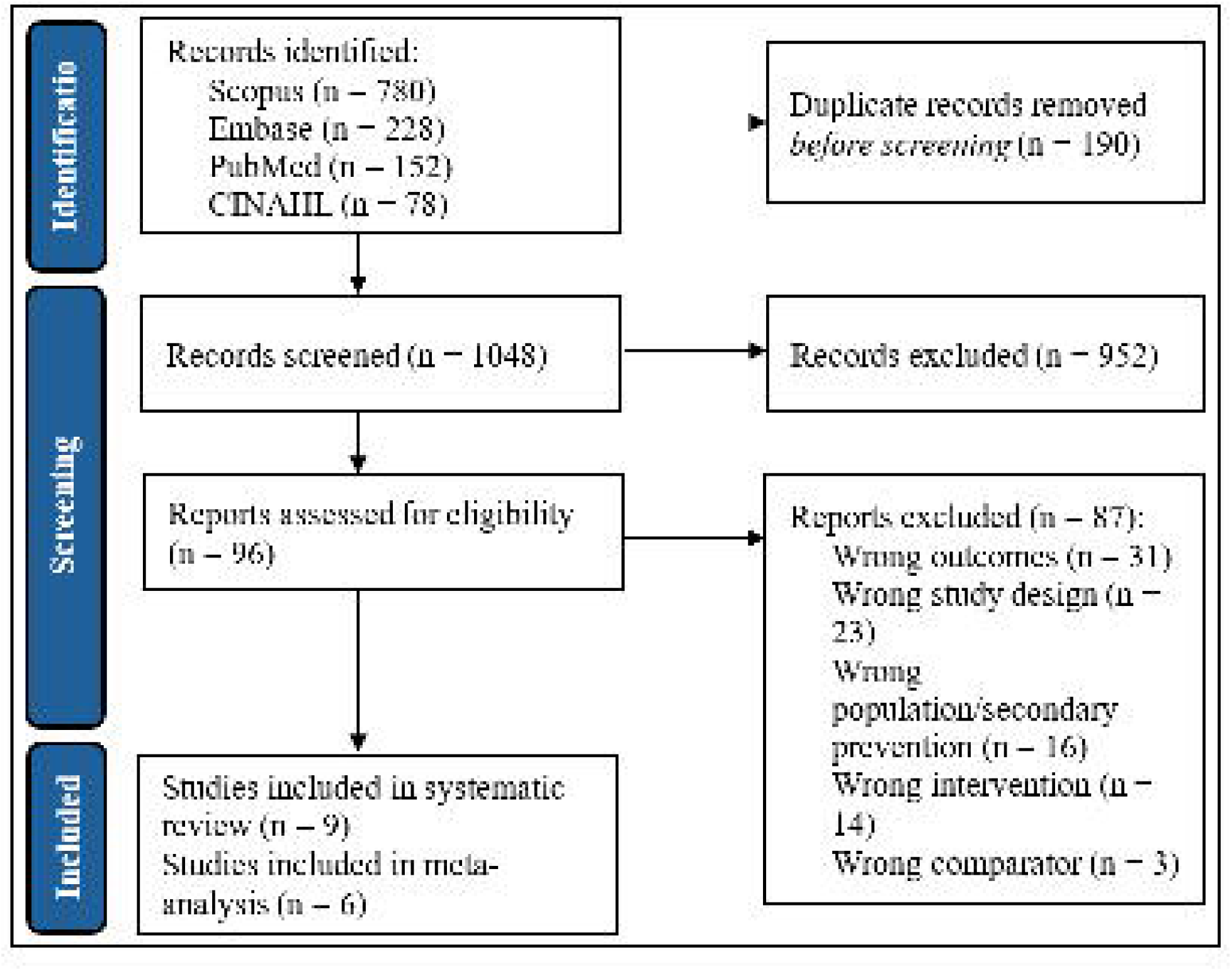
Preferred Reporting Items for Systematic Reviews and Meta-Analyses (PRISMA) flow diagram showing the study selection process

### 3.2 Study characteristics

This systematic review and meta-analysis included nine studies, the details of which are shown in **Table 1**. Four studies were conducted in North America (30, 31, 32, 33), five in Asia (31, 34, 35, 36, 37), two in Europe (31, 38), one in Australasia (31), one in South America (31), and one in Africa (31). Among these studies, one included participants from 32 countries spanning over six continents (31). The studies included RCTs (n=5) and NRSIs including intervention and cohort studies (n=4). Both the intervention and control groups included participants whose mean ages ranged from 52.2 years to approximately 68 years. To measure statin intolerance, six studies used discontinuation of statins (30, 31, 32, 34, 36, 38), five used adverse effects or events (30, 31, 34, 35, 36), three used muscle symptoms (30, 34, 35), three used myalgia (30, 31, 33), three used CK > 10 time the ULN or CK elevation (31, 37, 38), two used serious adverse events (30, 31), two used adverse drug reactions (30, 35), and one used statin intolerance (32). Of the nine included studies, three were not eligible for meta-analysis (32, 36, 38), because they either included the same population in the intervention and control groups or did not report no data on statin intolerance separately for the intervention and control group.

**Table 1:**
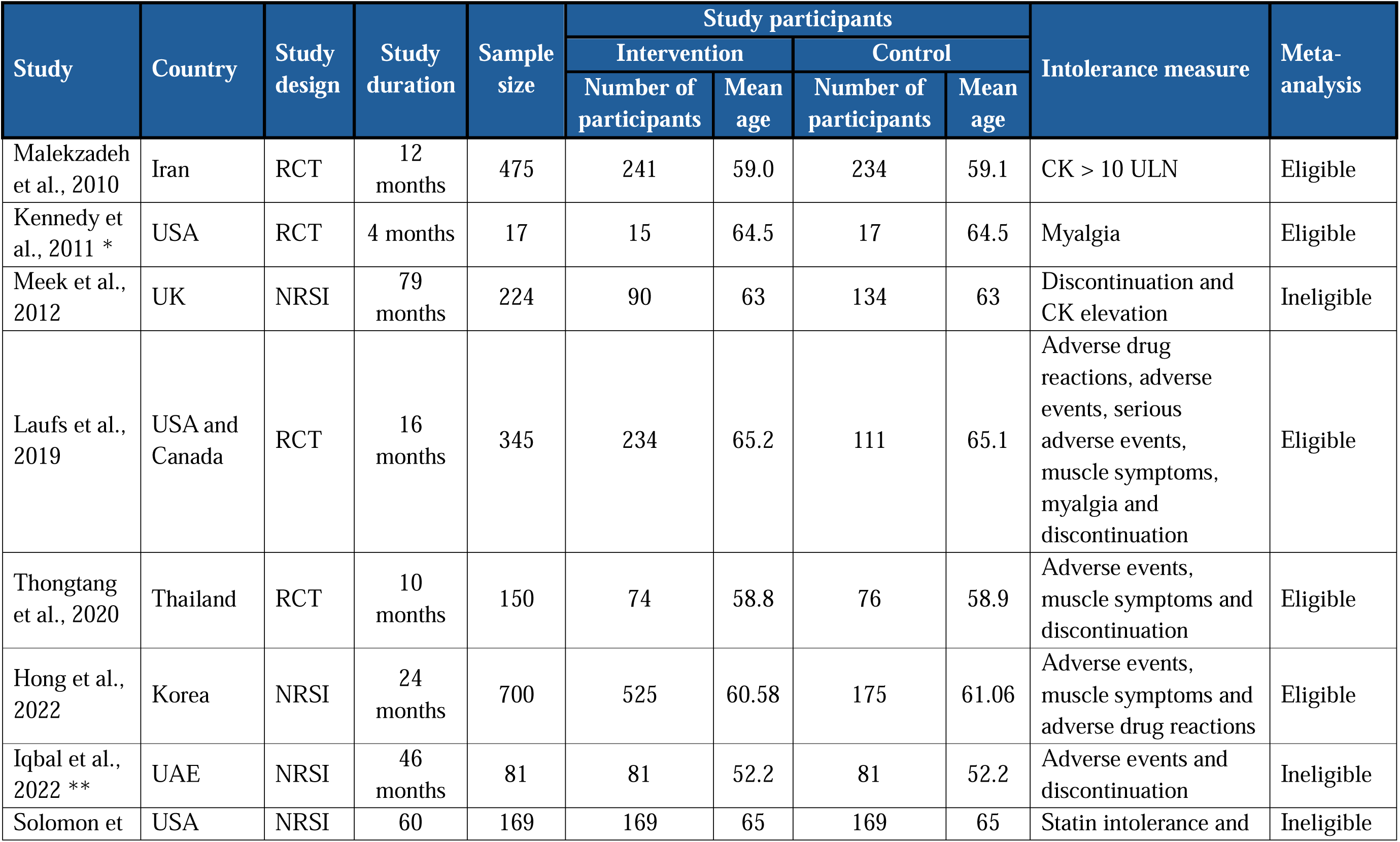
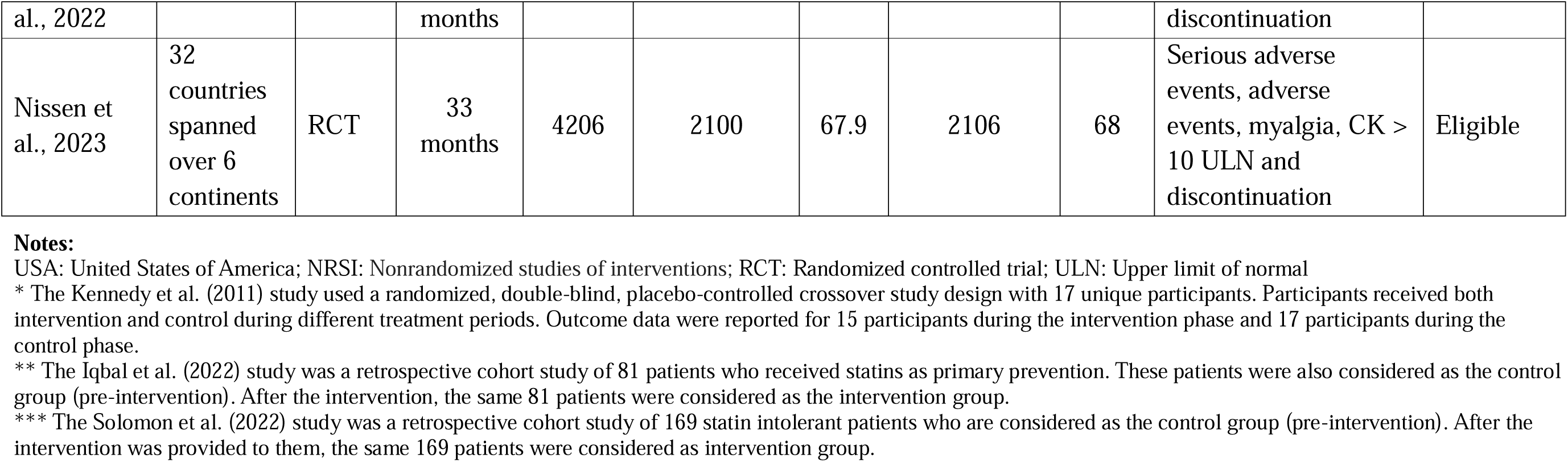
Characteristics of the included studies that assessed interventions to manage statin intolerance.

The risk of bias assessment of the nine included studies are shown in **Supplement 5**. Overall, approximately 22.2% of the studies (n=2) were high-risk, 66.7% (n=6) were of some concern, and 11.1% (n=1) were low-risk.

### 3.3 Intervention characteristics and key findings

**Table 2** shows that there were three adjuvant therapy interventions (35, 36, 37), including one multicomponent intervention that incorporated adjuvant therapy, three statin titration interventions (33, 34, 38), two interventions of replacing statins with other lipid-lowering agents (30, 31) and one intervention that involved switching to different statins (32). Three of the interventions took place in primary care settings (33, 35, 36), three took place in multicentre settings including primary care (30, 31, 38), one took place in a clinical trial setting (34), one took place in tertiary care setting (32) and the settings of one study was unclear (37). Three studies (32, 36, 37) included interventions delivered by general practitioners (GPs) and family doctors, and six studies (30, 31, 33, 34, 35, 38) did not specify the intervention deliverer. The objectives of all studies were to investigate or assess the tolerability or safety of an intervention in statin intolerant patients.

**Table 2:**
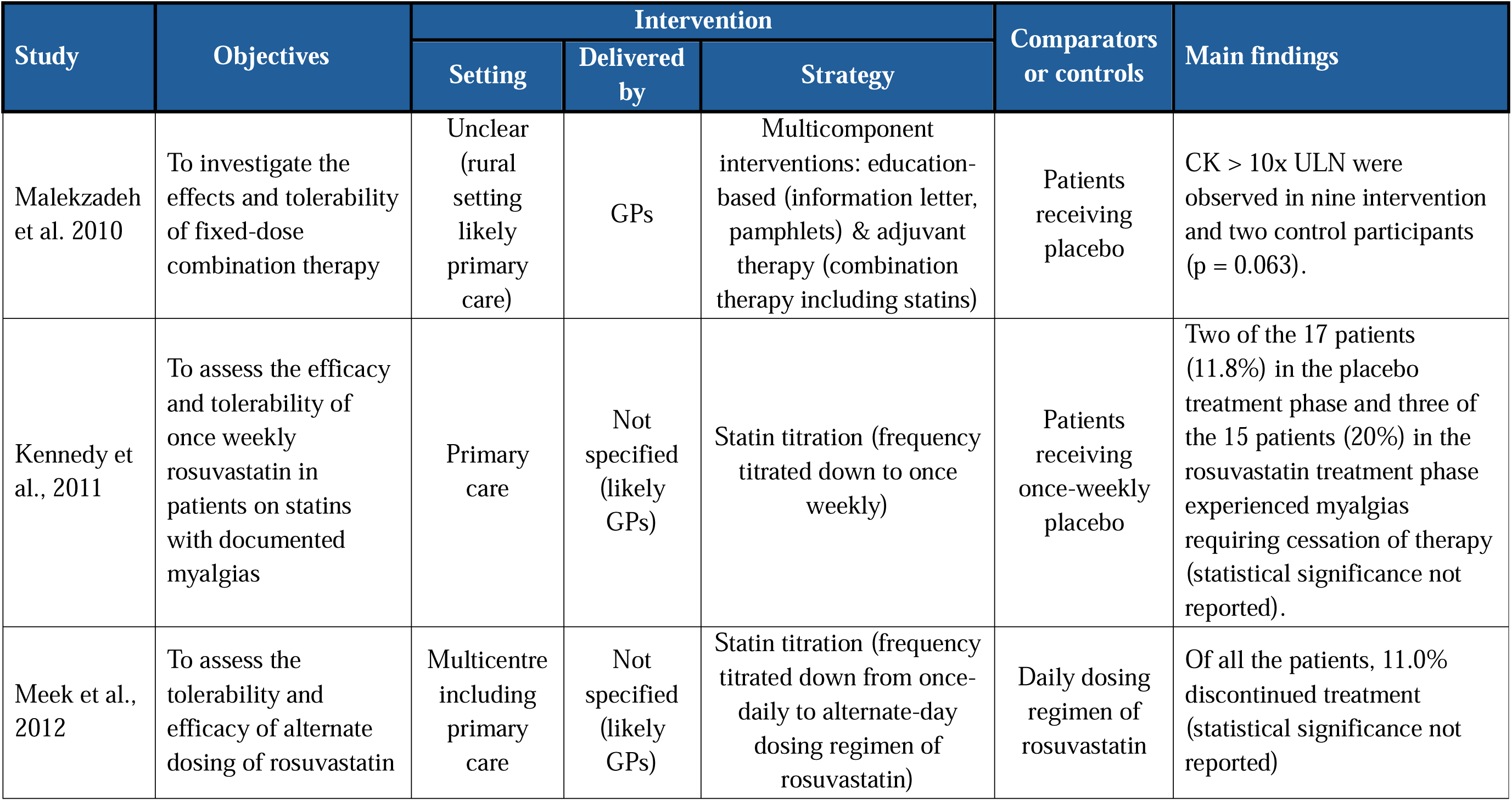
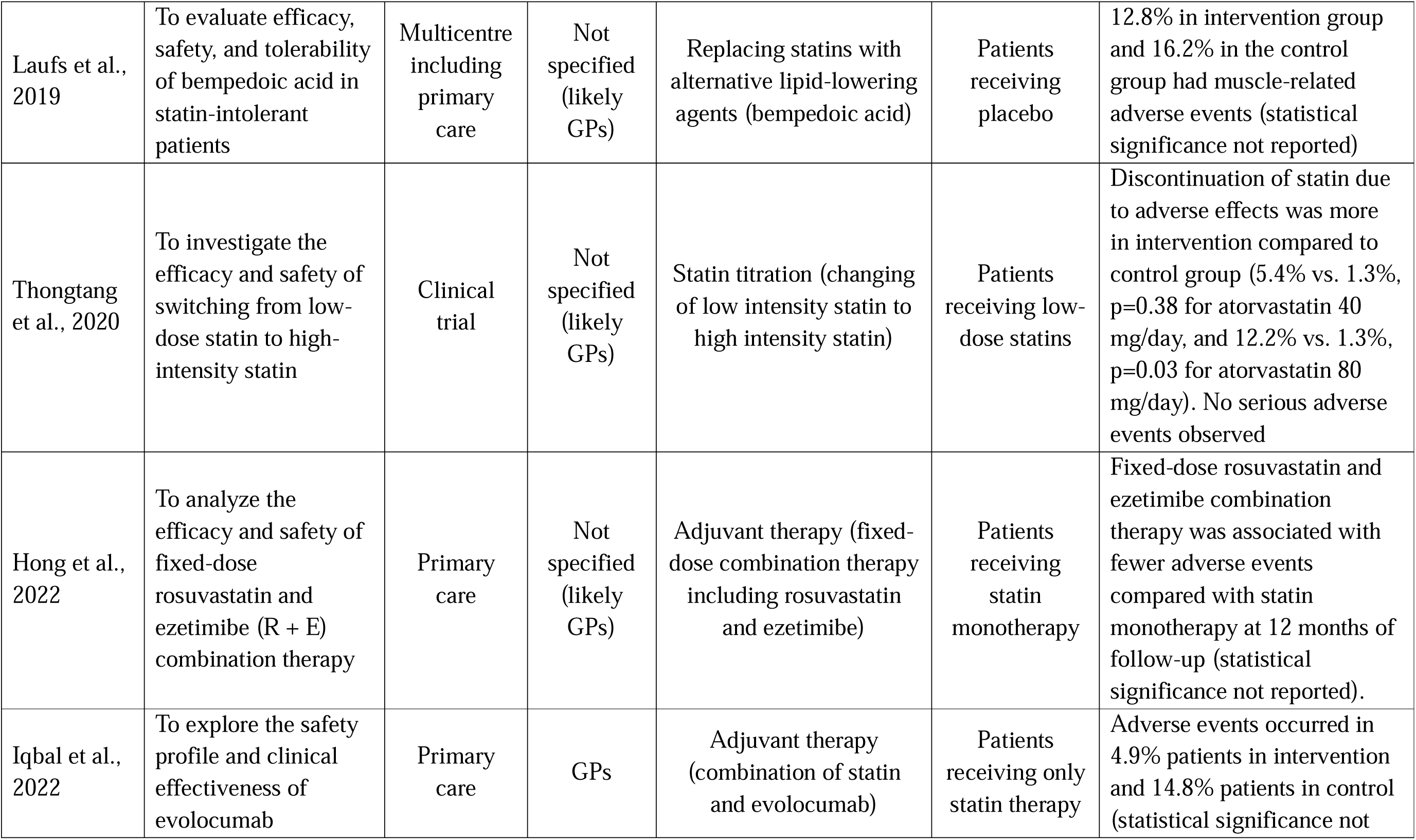
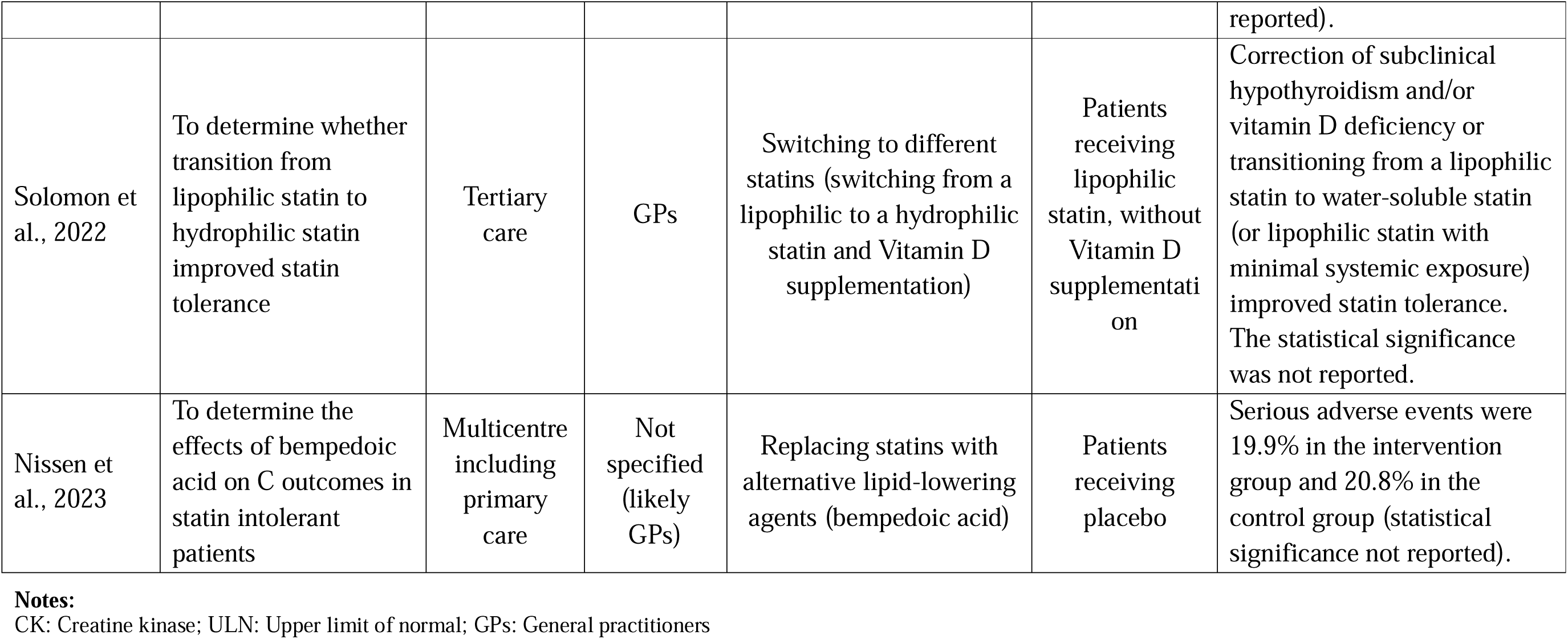
Characteristics of the interventions used and key findings of the included studies that assessed interventions to manage statin intolerance.

The key findings of this systematic review are summarised in **Table 2**. Overall, five studies (30, 31, 32, 35, 36) reported better outcomes for statin intolerance in the intervention group compared to the control group. Three studies (33, 34, 37) did not demonstrate better outcomes in the intervention group compared to the control group. One study (38) did not show any difference between the intervention group and the control group.

### 3.4 Reduction in adverse effects

#### Randomized controlled trials

Of the six eligible studies for meta-analysis, five were RCTs and one was an NRSI study. In the meta-analysis of these RCT studies that included 2,664 intervention participants and 2,544 control participants **(Figure 2a)**, the implemented interventions had a risk ratio (RR) of 0.97 (95% CI, 0.86–1.08), pooled across definitions of statin intolerance, relative to the control. The confidence interval (CI) included the possibility of no effect as it did not exclude 1.0. There was a moderate heterogeneity as depicted by an I-square of 27.4%.

**Figure 2:**
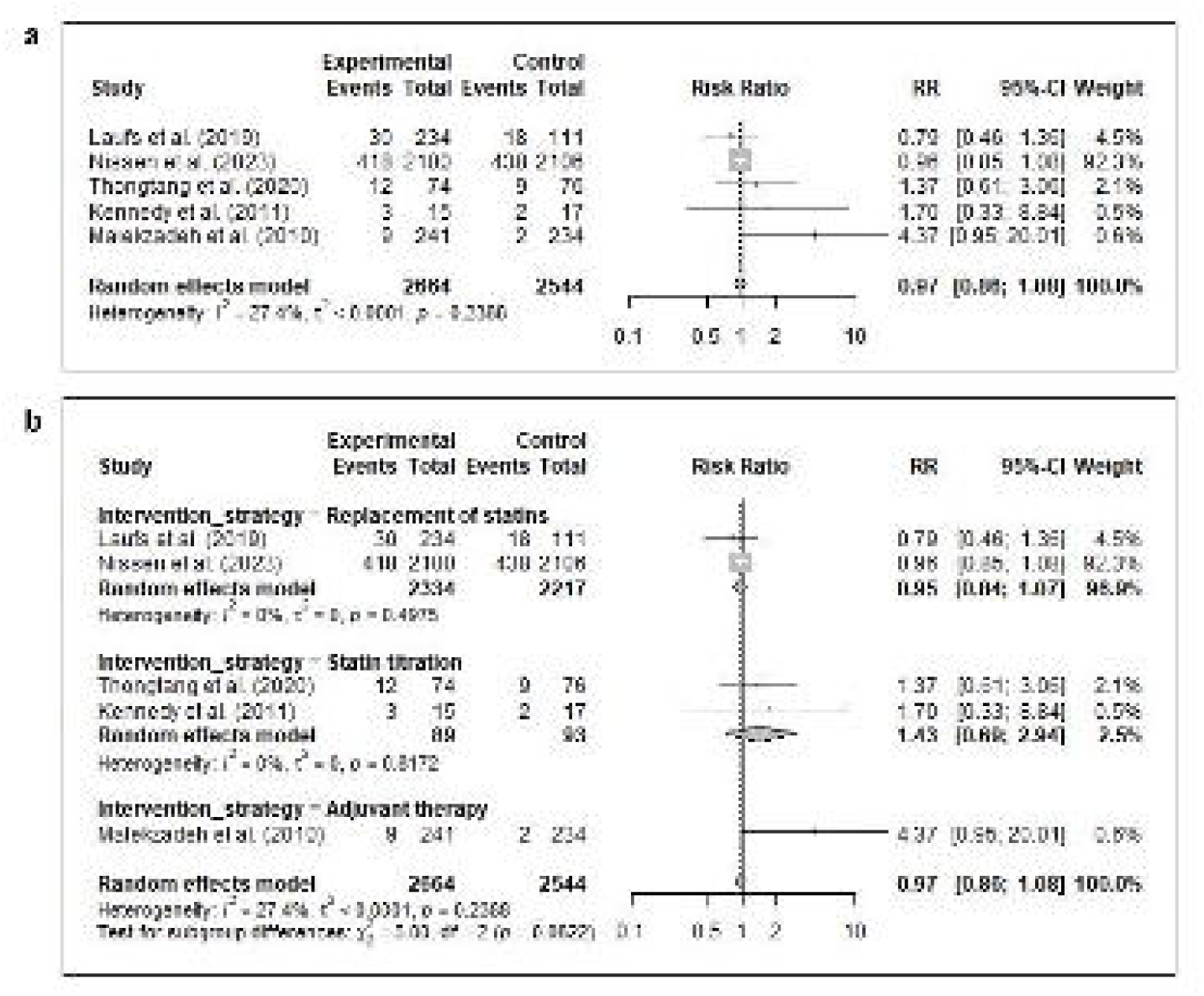
Forest plot for randomized controlled trials highlighting the effects of interventions on statin intolerance, as depicted by risk ratio **(a)** overall **(b)** in terms of various intervention strategies. Individual study estimates are shown as squares, with the size reflecting study weight, and horizontal lines represent 95% confidence intervals. Diamonds indicate pooled effect estimates derived using a random-effects model. An RR of 0.97 indicates 3% lower intolerance in the intervention group, while the 95% CI of 0.86–1.08 indicates weak confidence in the evidence. Overall, the plots demonstrate no reduction in intolerance with moderate degree of heterogeneity across studies. **Notes:** RR: Risk ratio; CI: Confidence interval

For the RCT studies, there was mixed evidence of an effect on statin intolerance for different intervention strategies relative to controls, and no significant evidence overall for a difference among subgroups (χ_2_^2^ = 5.0; p=0.08), as shown in **Figure 2b**. For interventions where statins were replaced with other lipid-lowering agents, the RR was 0.95 (95% CI, 0.84–1.07). For statin titration and adjuvant therapy, the RRs were 1.43 (95% CI, 0.69–2.94) and 4.37 (95% CI, 0.95–20.01), respectively. For all three intervention strategies, the CIs included the possibility of no effect as they did not exclude 1.0. There was no heterogeneity for all the intervention strategies. However, only one study included an adjuvant therapy intervention, two studies included replacing statins, and the rest of the two studies included statin titration. Due to the smaller number of studies in each subgroup, no clear conclusions can be drawn regarding whether the implemented interventions affect statin intolerance.

Further subgroup analyses were performed to determine whether participant age and study region were associated with statin intolerance in the RCT studies **(Supplement 6)**. For both studies in which the participants aged ≥65 years, the RR for statin intolerance relative to control was 0.95 (95% CI, 0.84–1.07). For participants aged <65 years, the RR was 1.75 (95% CI, 0.91–3.37). For both age subgroups, the CIs included the possibility of no effect as they did not exclude 1.0. In studies conducted in North America, those conducted across more than one region, and in the studies in Asia, the RRs were 0.85 (95% CI, 0.51–1.42), RR 0.96 (95% CI, 0.85–1.08), and 2.03 (95% CI, 0.69–5.95), respectively. The CIs for all three regional subgroups included the possibility of no effect as they did not exclude 1.0. Only one study was conducted in more than one region, and two studies conducted in each of North America and Asia. Due to the smaller number of studies in each subgroup, no clear conclusions can be drawn regarding how participant age and study region may affect statin intolerance.

#### Overall

The meta-analysis of all eligible studies, including the one NRSI study, showed a RR of 0.94, relative to control, pooled across definitions of statin intolerance **(Supplement 7)**. The 95% CI (0.63–1.42) crossed 1.0 in a substantial manner which meant that there was a high possibility of no effect. There was a substantial heterogeneity as depicted by an I-square of 56.4%. The results of the subgroup analyses including all eligible studies were consistent with those obtained from the subgroup analysis of the RCT studies. The details on all outcomes (such as muscle-related symptoms, myalgia, creatine kinase levels > 10x ULN) is shown in **Supplement 8**.

### 3.5 Statin discontinuation

Three studies (all RCTs) provided data on statin discontinuation. The meta-analysis of these studies including 2,408 intervention participants and 2,293 control participants showed a pooled RR of 0.99 (95% CI, 0.83–1.18) for statin discontinuation relative to the control, and the CI included the possibility of no effect as it did not exclude 1.0 **(Supplement 9)**. There was a low degree of heterogeneity as depicted by an I-square of 19.8%. However, due to the smaller number of studies, no clear conclusions can be drawn regarding how the implemented interventions affected statin discontinuation.

### 3.6 Publication bias

The risk of publication bias of studies on effects of interventions on statin intolerance was evaluated using a funnel plot **(Figure 3)** where the RR was compared with the standard error (SE) in the RR. Visual inspection suggested asymmetrical distributions, which may indicate publication bias and should be interpreted cautiously, particularly when the number of included studies is small.

**Figure 3:**
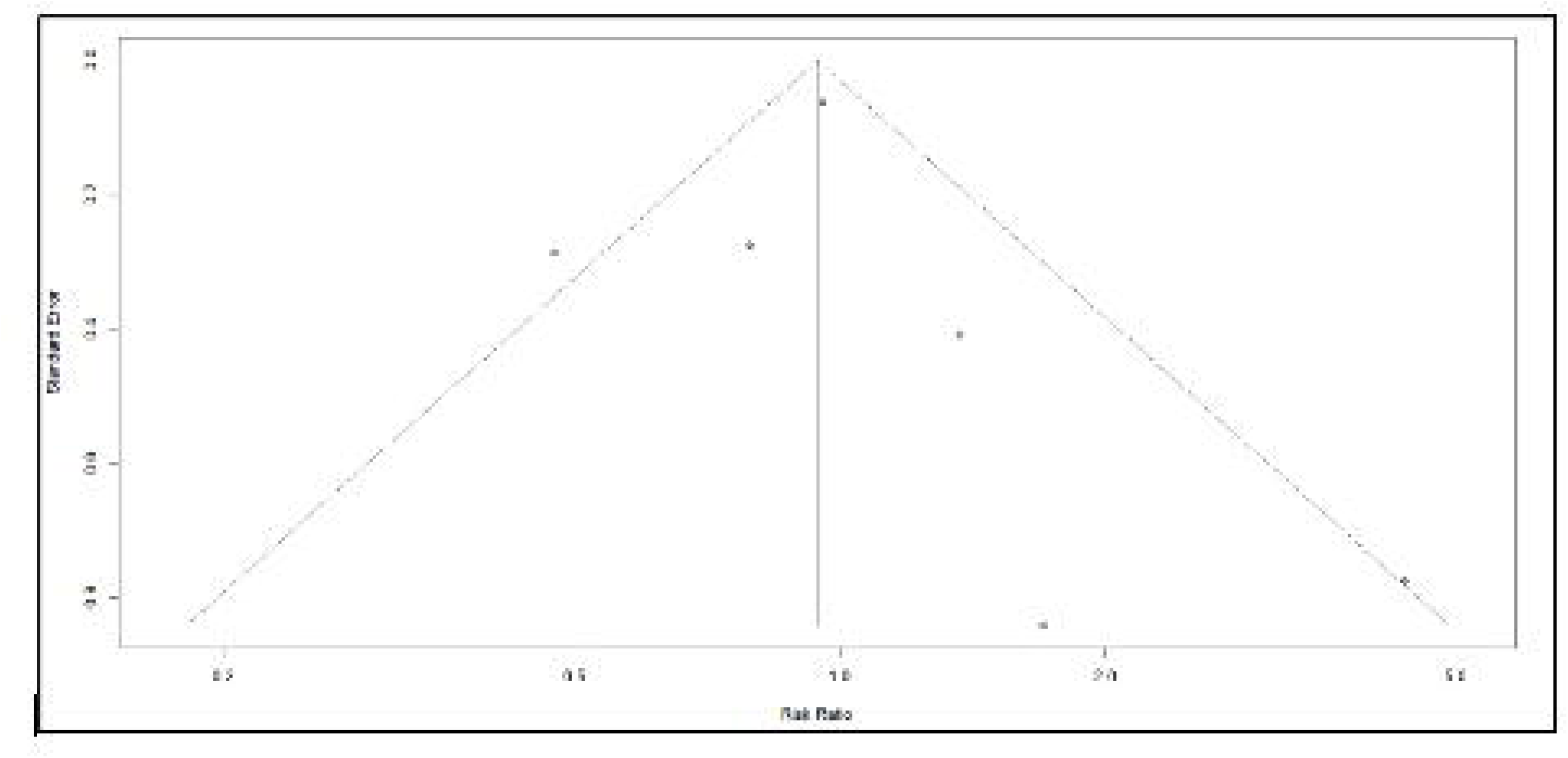
Publication bias of all studies on statin intolerance

## 4. Discussion

This systematic review found that, out of the nine included studies, interventions in five studies reduced statin intolerance in patients prescribed statins for primary prevention of CVD. Among the four studies in which the interventions did not reduce statin intolerance, three studies evaluated statin titration interventions (changing statin frequency or intensity) and one evaluated a multicomponent intervention consisting of adjuvant therapy (combination therapy including statins). One study did not show any difference between the intervention and control groups. All included studies aimed to investigate or assess the tolerability or safety of an intervention in statin intolerant patients. The pooled RR was 0.94 (95% CI, 0.63–1.42). The wide confidence interval indicated considerable imprecision, as the true effect could range from a meaningful reduction to a meaningful increase in intolerance. Several subgroup analyses were performed according to intervention strategies, mean participant ages, and study locations, which showed similar results. Overall, these findings do not provide strong evidence that any single intervention is consistently effective in reducing statin intolerance. This is broadly consistent with current guideline recommendations, which do not rely on a single management strategy but instead recommend an individualized and stepwise approach to suspected statin intolerance. This generally involves assessment of symptoms and other potential causes, followed where appropriate by temporary statin withdrawal and rechallenge, dose reduction, or switching to another statin after a washout period (39, 40). For patients who remain unable to tolerate an adequate statin regimen, non-statin lipid lowering therapies, including ezetimibe and PCSK9 inhibitors, may be considered according to the patient’s cardiovascular risk and the degree of cholesterol reduction required to achieve recommended treatment targets (24, 26). The findings of this review therefore do not support replacing this stepwise clinical approach with any one intervention evaluated in the included studies. Rather, they reinforce the need for management to be tailored to individual symptoms, treatment response, and cardiovascular risk (41, 42, 43).

Most of the six eligible studies for meta-analysis were conducted in primary care settings or multicentre settings that included primary care. Some studies specifically reported that the interventions were delivered by GPs, while the intervention provider was not specified in the remaining studies. These findings indicate that primary care was a common setting for the evaluated interventions, although the role of GPs in the intervention delivery could not be established across all included studies. Studies have shown that primary care physicians (PCPs) and GPs, who are usually the first points of contact for statin-intolerant patients, play important roles in the management of statin intolerance (39). Some studies show that electronic health records (EHRs) can be used by PCPs and GPs to identify and flag patients with muscle symptoms, myalgia, or adverse effects, potentially facilitating the delivery of targeted interventions to these patients (44). Furthermore, clinical settings are important for the management of statin intolerance, as clinicians can distinguish between patients that cannot tolerate statins at all and those that require alternative treatment options (15, 45, 46, 47). However, identifying statin intolerance using her data can be challenging because of the inconsistency and variability in the classification and definition of statin intolerance. The electronic phenotyping of statin intolerance is complex and require consideration of several characteristics, such as risk factors, diagnostic criteria, management strategies, symptom types, genetic predisposition, and pharmacological properties of statins (48, 49, 50). Therefore, a comprehensive understanding of the taxonomy of statin intolerance is crucial for the advancement of research on the identification and management of statin intolerance, for supporting CVD risk reduction.

This meta-analysis showed no strong evidence of reduced intolerance relative to control for participants aged ≥ 65 years. Statin intolerance is particularly challenging in older adults and the elderly because of age-related physiological changes, and the complexities of comorbidities and polypharmacy. However, the age at which interventions are most effective in managing statin intolerance requires further research. Studies have shown that tailored interventions may support the management of statin intolerance in patients aged ≥ 65 years (51, 52). Improved tolerance after statin rechallenge or dose alterations has also been reported among older populations (52, 53). Moreover, there are physiological (such as genetic) and psychological (such as nocebo effects) aspects of statin intolerance that could be considered when developing targeted interventions for older populations (51, 54, 55).

This meta-analysis did not provide any evidence that, compared to controls, the implemented interventions reduced statin discontinuation. This suggests that statin discontinuation is the cumulative effect of the intervention and participant characteristics. Interventions that manage statin intolerance may also improve statin adherence and reduce statin discontinuation, which would eventually improve patients’ health (56). Factors such as patient education and perceptions of statin-related adverse effects can influence the association between different intervention strategies and statin discontinuation, resulting in the inconclusive results of statin discontinuation in this review (9, 56). Studies show that patients experiencing statin-related adverse effects may exhibit low adherence rates or discontinuation (57). Patient-centred interventions may have a potential impact on statin adherence and discontinuation, although this effect may vary across patient demographics (58, 59).

Important evidence gaps nevertheless remain. The small number of studies, variation in definitions of statin intolerance and intervention strategies, and imprecision of the pooled estimates limit conclusions about which management approaches are most effective and for whom. Further studies using more consistent definitions of statin intolerance are needed to evaluate clinically relevant strategies such as rechallenge, dose modification, switching statins, non-statin lipid lowering therapies, and patient-centred interventions. Future research should also examine outcomes beyond intolerance alone, including statin adherence and persistence, treatment discontinuation, patient reported symptoms and experiences, and achievement of lipid targets. These studies would help determine how guideline recommended approaches can be translated into more consistent and effective management of statin intolerance in routine practice.

### Limitations

Most of the studies included in this systematic review and meta-analysis were assessed as having moderate risk of bias (or some concerns) to high risk of bias, reflecting issues such as incomplete outcome reporting and potential confounding in non-randomized designs. These limitations reduce the overall confidence in the pooled effect estimates and underscores the need for cautious interpretation. Limitations of this study also includes the increased risk of Type I error (finding false positives) due to the multiple subgroup analyses performed. Moreover, the funnel plot showed asymmetry, which may indicate publication bias. However, Egger’s test could not be performed to check for the presence of publication bias because the number of eligible studies in this meta-analysis did not meet the recommended number to test for asymmetry in funnel plots as defined by the Cochrane Handbook.

Another limitation of the study was that to narrow the search, the search syntax included only the term “primary prevention”, which may have resulted in the exclusion of studies that mentioned “primary and secondary prevention” **(Supplement 3.3)**. However, since these studies included both primary and secondary prevention patients, it may have been difficult to obtain data for only primary prevention patients. The variation in the definitions of statin intolerance, including adverse effects, statin-associated muscle symptoms and myalgia, also complicated the synthesis of findings across studies. The broader NLA definition of statin intolerance was therefore used in this study. However, a more specific definition was not within the scope of this study. Since there is currently no specific taxonomy for statin intolerance, and due to the limited number of eligible studies, it was not feasible to further stratify the studies according to specific definitions of statin intolerance, which limited the overall robustness of the findings of this study. Furthermore, due to the lack of a standardized taxonomy for statin intolerance, some of the eligible studies did not assess the more clinically relevant end point of the development of cardiovascular disease in patients who have undergone interventions for the management of statin intolerance (although this end point could be attributed to factors beyond statin intolerance, such as statin adherence). Finally, the included studies provided limited information on social factors such as education, and on the potential influence of nocebo/drucebo effects on reported statin intolerance. This limited the ability to examine the potential contribution of these factors to intervention outcomes.

## 5. Conclusion

This systematic review identified that the interventions used to manage statin intolerance in the primary prevention of CVD were adjuvant therapy, statin titration, replacing statins with other lipid-lowering agents, and switching to different statins. The meta-analysis found that among patients who were prescribed statins for primary prevention of CVD, the implemented interventions showed no strong evidence of reduced statin intolerance. The pooled RR was 0.97 (95% CI, 0.86–1.08) in RCT studies and 0.94 (95% CI, 0.63–1.42) in all studies, with both confidence intervals including the possibility of no effect as they did not exclude 1.0. Due to the small number of studies, no clear conclusions can be drawn regarding how the implemented interventions may affect statin discontinuation. As such, there is great uncertainty about the true effect of intervention strategies on statin intolerance. Clinicians might therefore consider this when providing interventions to patients to manage statin intolerance for primary prevention of CVD. Several steps can be taken to facilitate the understanding of statin intolerance and to provide guidance for clinicians to determine the best interventions to manage the condition and to improve statin adherence which are fundamental barriers to improving the overall health outcomes of patients at risk of CVD. Most importantly, there is a need to develop a standardized taxonomy to better define and identify patients with statin intolerance. Furthermore, there is also a need to develop and establish an electronic phenotyping algorithm to identify statin intolerant patients from electronic health records. In summary, there is a clear need to focus more on research related to the role of general practitioners in delivering patient-centred interventions to manage statin intolerance in patients receiving statins for primary prevention of CVD.

## Data Availability

The corresponding author will make the data available upon request.

## Author contributions

1. SR and JJ contributed to the conceptualization of this study.
2. JJ contributed to the funding acquisition
3. JR, KAR, STL and JJ contributed to the resources and supervision of this study
4. JR, KAR, STL and JJ contributed to the validation
5. All authors contributed to the reviewing and editing of this study
6. SR and JJ contributed to data curation
7. SR contributed to the formal analysis and the original drafting of this study
8. SR contributed to the methodology of the study

## Data sharing statement

The corresponding author will make the data available upon request.

## Acknowledgements

We would like to thank the ePBRN Primary Care Health Informatics Working Group of the Secure Research Environment for Digital Health (SREDH) Consortium (www.sredhconsortium.org, accessed on 7 October 2024) for their assistance with access to the ePBRN dataset to investigate the findings from this review.

