## Supplement for "Interventions to manage intolerance among patients prescribed statins for primary prevention of cardiovascular diseases: A systematic review and meta-analysis"

**Table of contents**

**Supplement 1: PRISMA 2020 checklist ....…………………………….………..…….…... 2**

**Supplement 2: Table showing the inclusion and exclusion criteria ...………….….…...... 8**

**Supplement 3: Tables for search syntax …………………..……….………………….… 10**

Supplement 3A: Search syntax in PubMed ….……………….……………………….……. 10

Supplement 3B: Search syntax in Scopus ………………….…………………………….… 12

Supplement 3C: Search syntax in Ovid ……………….…….….…………........................... 13

Supplement 3D: Search syntax in CINAHL ….………………………………………….… 15

**Supplement 4: List of excluded studies …….…………..………………………………... 18**

**Supplement 5: Risk of bias and quality assessment of included studies ………………. 28**

Supplement 5A: Quality assessment of randomized controlled trials using the RoB 2 tool....28

Supplement 5B: Quality assessment of nonrandomized studies of interventions using the ROBINS-I tool …………………..………………………………………………………..... 29

**Supplement 6: Subgroup analysis (forest plots of RCTs) for statin intolerance ............. 30**

Supplement 6A: Role of participant age in the effectiveness of intervention …………….... 30

Supplement 6B: Role of geographic location in the effectiveness of intervention ……….... 31

**Supplement 7: Subgroup analysis (forest plots of all studies including RCTs and NRSIs) for statin intolerance .….…..……………………………………………….……………... 32**

Supplement 7A: Role of intervention strategy in the effectiveness of intervention ……….. 32

Supplement 7B: Role of participant age in the effectiveness of intervention …………….... 33

Supplement 7C: Role of geographic location in the effectiveness of intervention ..……….. 34

**Supplement 8: Details of all outcomes ….………………………………….…………….. 35**

**Supplement 9: Subgroup analysis (forest plots) for statin discontinuation ………….... 36**

**Supplement 1: PRISMA 2020 checklist**

| **Section and Topic** | **Item #** | **Checklist item** | **Location where item is reported** |
| --- | --- | --- | --- |
| **TITLE** | | |  |
| Title | 1 | Identify the report as a systematic review. | Page 1 |
| **ABSTRACT** | | |  |
| Abstract | 2 | See the PRISMA 2020 for Abstracts checklist. | Page 2 (Abstract) |
| **INTRODUCTION** | | |  |
| Rationale | 3 | Describe the rationale for the review in the context of existing knowledge. | Pages 3-4 (Introduction) |
| Objectives | 4 | Provide an explicit statement of the objective(s) or question(s) the review addresses. | Page 4 |
| **METHODS** | | |  |
| Eligibility criteria | 5 | Specify the inclusion and exclusion criteria for the review and how studies were grouped for the syntheses. | - Page 4 (Eligibility criteria) - Supplement 2 |
| Information sources | 6 | Specify all databases, registers, websites, organisations, reference lists and other sources searched or consulted to identify studies. Specify the date when each source was last searched or consulted. | - Page 5 (Search strategy) - Supplement 3 |
| Search strategy | 7 | Present the full search strategies for all databases, registers and websites, including any filters and limits used. | - Supplement 3 - Page 5 (Search strategy) |
| Selection process | 8 | Specify the methods used to decide whether a study met the inclusion criteria of the review, including how many reviewers screened each record and each report retrieved, whether they worked independently, and if applicable, details of automation tools used in the process. | Page 5 (Data collection) |
| Data collection process | 9 | Specify the methods used to collect data from reports, including how many reviewers collected data from each report, whether they worked independently, any processes for obtaining or confirming data from study investigators, and if applicable, details of automation tools used in the process. | Page 5 (Data collection) |
| Data items | 10a | List and define all outcomes for which data were sought. Specify whether all results that were compatible with each outcome domain in each study were sought (e.g. for all measures, time points, analyses), and if not, the methods used to decide which results to collect. | - Pages 6-7 (Data synthesis and analysis) - PROSPERO protocol – CRD42024587573 |
|  | 10b | List and define all other variables for which data were sought (e.g. participant and intervention characteristics, funding sources). Describe any assumptions made about any missing or unclear information. | - Page 5 (Data collection) - PROSPERO protocol – CRD42024587573 |
| Study risk of bias assessment | 11 | Specify the methods used to assess risk of bias in the included studies, including details of the tool(s) used, how many reviewers assessed each study and whether they worked independently, and if applicable, details of automation tools used in the process. | Pages 5-6 (Risk of bias assessment) |
| Effect measures | 12 | Specify for each outcome the effect measure(s) (e.g. risk ratio, mean difference) used in the synthesis or presentation of results. | Pages 6-7 (Data synthesis and analysis) |
| Synthesis methods | 13a | Describe the processes used to decide which studies were eligible for each synthesis (e.g. tabulating the study intervention characteristics and comparing against the planned groups for each synthesis (item #5)). | - Pages 6-7 (Data synthesis and analysis) - PROSPERO protocol – CRD42024587573 |
|  | 13b | Describe any methods required to prepare the data for presentation or synthesis, such as handling of missing summary statistics, or data conversions. | - Pages 6-7 (Data synthesis and analysis) - PROSPERO protocol – CRD42024587573 |
|  | 13c | Describe any methods used to tabulate or visually display results of individual studies and syntheses. | - Pages 6-7 (Data synthesis and analysis) - PROSPERO protocol – CRD42024587573 |
|  | 13d | Describe any methods used to synthesize results and provide a rationale for the choice(s). If meta-analysis was performed, describe the model(s), method(s) to identify the presence and extent of statistical heterogeneity, and software package(s) used. | - Pages 6-7 (Data synthesis and analysis) - PROSPERO protocol – CRD42024587573 |
|  | 13e | Describe any methods used to explore possible causes of heterogeneity among study results (e.g. subgroup analysis, meta-regression). | - Pages 6-7 (Data synthesis and analysis) - PROSPERO protocol – CRD42024587573 |
|  | 13f | Describe any sensitivity analyses conducted to assess robustness of the synthesized results. | - Pages 6-7 (Data synthesis and analysis) - PROSPERO protocol – CRD42024587573 |
| Reporting bias assessment | 14 | Describe any methods used to assess risk of bias due to missing results in a synthesis (arising from reporting biases). | Pages 5-6 (Risk of bias assessment) |
| Certainty assessment | 15 | Describe any methods used to assess certainty (or confidence) in the body of evidence for an outcome. | - Pages 6-7 (Data synthesis and analysis) - PROSPERO protocol – CRD42024587573 |
| **RESULTS** | | |  |
| Study selection | 16a | Describe the results of the search and selection process, from the number of records identified in the search to the number of studies included in the review, ideally using a flow diagram. | - Page 7 (Selection of studies) - Figure 1 |
|  | 16b | Cite studies that might appear to meet the inclusion criteria, but which were excluded, and explain why they were excluded. | Supplement 4 |
| Study characteristics | 17 | Cite each included study and present its characteristics. | Pages 8-10 (Study characteristics) |
| Risk of bias in studies | 18 | Present assessments of risk of bias for each included study. | - Pages 8-10 (Study characteristics) - Supplement 5 |
| Results of individual studies | 19 | For all outcomes, present, for each study: (a) summary statistics for each group (where appropriate) and (b) an effect estimate and its precision (e.g. confidence/credible interval), ideally using structured tables or plots. | Pages 11-14 (Intervention characteristics and key findings) |
| Results of syntheses | 20a | For each synthesis, briefly summarise the characteristics and risk of bias among contributing studies. | - Pages 8-10 (Study characteristics) - Pages 11-14 (Intervention characteristics and key findings) - Supplement 5 |
|  | 20b | Present results of all statistical syntheses conducted. If meta-analysis was done, present for each the summary estimate and its precision (e.g. confidence/credible interval) and measures of statistical heterogeneity. If comparing groups, describe the direction of the effect. | - Pages 156-158 (Reduction in adverse effects) - Page 158 (Statin discontinuation) - Figure 2 - Supplement 6 - Supplement 7 - Supplement 8 |
|  | 20c | Present results of all investigations of possible causes of heterogeneity among study results. | - Pages 15-17 (Reduction in adverse effects) - Page 17 (Statin discontinuation) - Figure 2 - Supplement 6 - Supplement 7 - Supplement 8 |
|  | 20d | Present results of all sensitivity analyses conducted to assess the robustness of the synthesized results. | - Pages 15-17 (Reduction in adverse effects) - Page 17 (Statin discontinuation) - Figure 2 - Supplement 6 - Supplement 7 - Supplement 8 |
| Reporting biases | 21 | Present assessments of risk of bias due to missing results (arising from reporting biases) for each synthesis assessed. | - Pages 17-18 (Publication bias) - Figure 3 |
| Certainty of evidence | 22 | Present assessments of certainty (or confidence) in the body of evidence for each outcome assessed. | - Pages 17-18 (Publication bias) - Figure 3 |
| **DISCUSSION** | | |  |
| Discussion | 23a | Provide a general interpretation of the results in the context of other evidence. | Pages 18-20 (Discussion) |
|  | 23b | Discuss any limitations of the evidence included in the review. | Pages 20-21 (Limitations) |
|  | 23c | Discuss any limitations of the review processes used. | Pages 20-21 (Limitations) |
|  | 23d | Discuss implications of the results for practice, policy, and future research. | Pages 22 (Conclusions) |
| **OTHER INFORMATION** | | |  |
| Registration and protocol | 24a | Provide registration information for the review, including register name and registration number, or state that the review was not registered. | Page 4 (Methods) |
|  | 24b | Indicate where the review protocol can be accessed, or state that a protocol was not prepared. | Page 4 (Methods) |
|  | 24c | Describe and explain any amendments to information provided at registration or in the protocol. | Page 4 (Methods) |
| Support | 25 | Describe sources of financial or non-financial support for the review, and the role of the funders or sponsors in the review. | Page 23 |
| Competing interests | 26 | Declare any competing interests of review authors. | Page 23 |
| Availability of data, code and other materials | 27 | Report which of the following are publicly available and where they can be found: template data collection forms; data extracted from included studies; data used for all analyses; analytic code; any other materials used in the review. | Page 23 |

*From:* Page MJ, McKenzie JE, Bossuyt PM, Boutron I, Hoffmann TC, Mulrow CD, et al. The PRISMA 2020 statement: an updated guideline for reporting systematic reviews. BMJ 2021;372:n71. doi: 10.1136/bmj.n71. This work is licensed under CC BY 4.0. To view a copy of this license, visit <https://creativecommons.org/licenses/by/4.0/>

**Supplement 2: Table showing the inclusion and exclusion criteria**

| **PICOS** | **Inclusion criteria** | **Exclusion criteria** |
| --- | --- | --- |
| **Population** | - Adult participants aged ≥ 18 years - People who are prescribed statins for primary prevention at a primary healthcare facility - Population of all ethnicity, gender, and geographical location | - Participants aged <18 years - People who are prescribed statins for secondary prevention |
| **Interventions** | - Interventions that address statin intolerance - Interventions that target patients for primary prevention of CVD - Interventions that target specific groups (such as patients with Alzheimer’s Disease or a prison population), inpatients (those admitted at health facilities/hospitals), or outpatients (those who receive a statin prescription without being admitted at any health facilities/hospitals) - Interventions that target people who are prescribed only a statin or prescribed multiple medications including a statin - Interventions that are general practitioners (GP)-led, prescribers-led, pharmacist-led, family physician-led - Among others, interventions can be combination therapies including statins, changing of statin type, changing of statin dose and/or intensity, changing to or adding other lipid-lowering drugs, or multicomponent | Interventions that target patients who have already been deprescribed for statins or no longer takes statins. |
| **Controls or comparators** | Controls that include either placebo, usual care (no change in the usual activities of care or no intervention), or statin monotherapy | Studies with no control group (placebo, usual care, or monotherapy) |
| **Outcomes** | Primary outcome of interest is any data or statistics related to tolerance outcomes. This will include number patients with adverse events, adverse effect, serious adverse events, muscle related symptoms, myalgia, creatine kinase levels > 10 ULN | Studies that do not report data or statistics related to tolerance outcomes |
| **Studies** | - Studies including electronic health record (EHR) and non-EHR data - Clinical studies, trials, intervention studies (experimental studies, randomized controlled trials), and any cohort study investigating any intervention, strategy, or program that addressed statin intolerance - Studies published in peer-reviewed journals until June 2025 - Studies published in English language | Qualitative studies, simulation modeling studies, protocols, feasibility data, opinion literatures, commentaries, editorials, brief reports, prospectives, conference abstracts, proceedings, thesis, animal model studies, observational studies (case‒control studies, cross-sectional studies), reviews, systematic reviews and meta-analysis |

**Supplement 3: Tables for search syntax**

| **Keyword groups** | **Search Syntax in PubMed** |
| --- | --- |
| **Primary care** | 1. primary care[MeSH Terms] 2. primary care[Title/Abstract] 3. primary healthcare[Title/Abstract] 4. primary medical care[Title/Abstract] 5. primary prevention[Title/Abstract] 6. general practice[Title/Abstract] 7. general practitioner[Title/Abstract] 8. community pharmacy[Title/Abstract] 9. rural healthcare[Title/Abstract] 10. district healthcare[Title/Abstract] |
| **Disease condition** | 11. cardiovascular diseases[MeSH Terms] 12. hyperlipidemia[MeSH Terms] 13. dyslipidemia[MeSH Terms] 14. hypertension[Title/Abstract] 15. cardiovascular disease[Title/Abstract] 16. CVD[Title/Abstract] 17. Cardiovascular disease risk factor[Title/Abstract] 18. Angina[Title/Abstract] 19. CVD risk factor [Title/Abstract] 20. atherosclerotic cardiovascular disease risk factor[Title/Abstract] 21. dyslipidemia[Title/Abstract] 22. hyperlipidemia[Title/Abstract] 23. Hyperlipid[Title/Abstract] 24. Hypercholesterolemia[Title/Abstract] 25. Hypercholesterol[Title/Abstract] 26. Familial hyperlipidemia[Title/Abstract] 27. heart disease[Title/Abstract] 28. atherosclerotic disease risk factor[Title/Abstract] 29. cardiovascular risk[Title/Abstract] 30. cholesterolemia[Title/Abstract] 31. cardiovascular[Title/Abstract] 32. cholesterol[Title/Abstract] 33. High blood pressure[Title/Abstract] 34. Overweight[Title/Abstract] 35. Obese[Title/Abstract] 36. Obesity[Title/Abstract] |

**Supplement 3A: Search syntax in PubMed**

| **Statin** | 37. Hydroxymethylglutaryl‐CoA Reductase Inhibitors[MeSH Terms] 38. statin[MeSH Terms] 39. Hydroxymethylglutaryl‐CoA Reductase Inhibitors[Title/Abstract] 40. statin[Title/Abstract] 41. statins[Title/Abstract] 42. *statin[Title/Abstract] 43. statin prescribing[Title/Abstract] 44. lipid lowering[Title/Abstract] 45. cholesterol lowering[Title/Abstract] 46. lipitor[Title/Abstract] 47. baycol[Title/Abstract] 48. lescol[Title/Abstract] 49. mevacor[Title/Abstract] 50. altocor[Title/Abstract] 51. pravachol[Title/Abstract] 52. lipostat[Title/Abstract] 53. zocor[Title/Abstract] 54. mevinolin[Title/Abstract] 55. compactin[Title/Abstract] 56. altoprev[Title/Abstract] 57. zypitamag[Title/Abstract] 58. ezallor[Title/Abstract] |
| --- | --- |
| **Tolerance** | 59. Drug Tolerance[MeSH Terms] 60. tolerance[Title/Abstract] 61. intolerance[Title/Abstract] 62. tolerant[Title/Abstract] 63. intolerant[Title/Abstract] 64. tolerability[Title/Abstract] 65. intolerability[Title/Abstract] |
| **Combinations** | 66. #1 OR #2 OR #3 OR #4 OR #5 OR #6 OR #7 OR #8 OR #9 OR #10 67. #11 OR #12 OR #13 OR #14 OR #15 OR #16 OR #17 OR #18 OR #19 OR #20 OR #21 OR #22 OR #23 OR #24 OR #25 OR #26 OR #27 OR #28 OR #29 OR #30 OR #31 OR #32 OR #33 OR #34 OR #35 OR #36 68. #37 OR #38 OR #39 OR #40 OR #41 OR #42 OR #43 OR #44 OR #45 OR #46 OR #47 OR #48 OR #49 OR #50 OR #51 OR #52 OR #53 OR #54 OR #55 OR #56 OR #57 OR #58 69. #59 OR #60 OR #61 OR #62 OR #63 OR #64 OR #65 70. #66 AND #67 AND #68 AND #69 71. #66 AND #67 AND #68 AND #69 |

**Supplement 3B: Search syntax in Scopus**

| **Keyword groups** | **Search Syntax in Scopus** |
| --- | --- |
| **Primary care** | 1. TITLE-ABS-KEY ("primary care" OR "primary healthcare" OR "primary medical care" OR "primary prevention" OR "general practice" OR "general practitioner" OR "community pharmacy" OR "rural healthcare" OR "district healthcare") |
| **Disease condition** | 2. TITLE-ABS-KEY ("cardiovascular diseases" OR "hypertension" OR "cardiovascular disease" OR "CVD" OR "Cardiovascular disease risk factor" OR "Angina" OR "CVD risk factor" OR "atherosclerotic cardiovascular disease risk factor" OR "dyslipidemia" OR "hyperlipidemia" OR "Hyperlipid" OR "Hypercholesterolemia" OR "Hypercholesterol" OR "Familial hyperlipidemia" OR "heart disease" OR "atherosclerotic disease risk factor" OR "cardiovascular risk" OR "cholesterolemia" OR "cardiovascular" OR "cholesterol" OR "High blood pressure" OR "Overweight" OR "Obese" OR "Obesity" ) |
| **Statin** | 3. TITLE-ABS-KEY ("Hydroxymethylglutaryl‐CoA Reductase Inhibitors" OR "statin" OR "statins" OR "*statin" OR "statin prescribing" OR "lipid lowering" OR "cholesterol lowering" OR "Lipitor" OR "baycol" OR "lescol" OR "mevacor" OR "altocor" OR "Pravachol" OR "lipostat" OR "zocor" OR "Mevinolin" OR "compactin" OR "altoprev" OR "zypitamag" OR "ezallor" ) |
| **Tolerance** | 4. TITLE-ABS-KEY (" tolerance" OR " intolerance" OR "tolerant" OR "intolerant" OR "tolerability" OR "intolerability") |
| **Combinations (2013 to 2024)** | 5. (TITLE-ABS-KEY ("primary care" OR "primary healthcare" OR "primary medical care" OR "primary prevention" OR "general practice" OR "general practitioner" OR "community pharmacy" OR "rural healthcare" OR "district healthcare" ) AND TITLE-ABS-KEY ("cardiovascular diseases" OR "hypertension" OR "cardiovascular disease" OR "CVD" OR "Cardiovascular disease risk factor" OR "Angina" OR "CVD risk factor" OR "atherosclerotic cardiovascular disease risk factor" OR "dyslipidemia" OR "hyperlipidemia" OR "Hyperlipid" OR "Hypercholesterolemia" OR "Hypercholesterol" OR "Familial hyperlipidemia" OR "heart disease" OR "atherosclerotic disease risk factor" OR "cardiovascular risk" OR "cholesterolemia" OR "cardiovascular" OR "cholesterol" OR "High blood pressure" OR "Overweight" OR "Obese" OR "Obesity" ) AND TITLE-ABS-KEY ("Hydroxymethylglutaryl‐CoA Reductase Inhibitors" OR "statin" OR "statins" OR "*statin" OR "statin prescribing" OR "lipid lowering" OR "cholesterol lowering" OR "Lipitor" OR "baycol" OR "lescol" OR "mevacor" OR "altocor" OR "Pravachol" OR "lipostat" OR "zocor" OR "Mevinolin" OR "compactin" OR "altoprev" OR "zypitamag" OR "ezallor" ) AND TITLE-ABS-KEY (" tolerance" OR " intolerance" OR "tolerant" OR "intolerant" OR "tolerability" OR "intolerability" ) ) |

**Supplement 3C: Search syntax in Ovid**

| **Keyword groups** | **Search Syntax in Embase Ovid** |
| --- | --- |
| **Primary care** | 1. primary medical care/ 2. primary health care/ 3. primary healthcare.ti, ab, kw. 4. primary care.ti, ab, kw. 5. primary medical care.ti, ab, kw. 6. primary prevention.ti, ab, kw. 7. general practice.ti, ab, kw. 8. general practitioner.ti, ab, kw. 9. community pharmacy.ti, ab, kw. 10. rural healthcare.ti, ab, kw. 11. district healthcare.ti, ab, kw. |
| **Disease condition** | 12. cardiovascular disease/ 13. hyperlipidemia/ 14. dyslipidemia/ 15. hypertension.ti, ab, kw. 16. cardiovascular disease.ti, ab, kw. 17. CVD.ti, ab, kw. 18. Cardiovascular disease risk factor.ti, ab, kw. 19. Angina.ti, ab, kw. 20. CVD risk factor.ti, ab, kw. 21. atherosclerotic cardiovascular disease risk factor.ti, ab, kw. 22. dyslipidemia.ti, ab, kw. 23. hyperlipidemia.ti, ab, kw. 24. Hyperlipid.ti, ab, kw. 25. Hypercholesterolemia.ti, ab, kw. 26. Hypercholesterol.ti, ab, kw. 27. Familial hyperlipidemia.ti, ab, kw. 28. heart disease.ti, ab, kw. 29. atherosclerotic disease risk factor.ti, ab, kw. 30. cardiovascular risk.ti, ab, kw. 31. cholesterolemia.ti, ab, kw. 32. cardiovascular.ti, ab, kw. 33. cholesterol.ti, ab, kw. 34. High blood pressure.ti, ab, kw. 35. Overweight.ti, ab, kw. 36. Obese.ti, ab, kw. 37. Obesity.ti, ab, kw. |
| **Statin** | 38. hydroxymethylglutaryl coenzyme A reductase inhibitor/ 39. statin.ti, ab, kw. 40. statins.ti, ab, kw. 41. ?statin.ti, ab, kw. 42. statin prescribing.ti, ab, kw. 43. lipid lowering.ti, ab, kw. 44. cholesterol lowering.ti, ab, kw. |
|  | 45. lipitor.ti, ab, kw. 46. baycol.ti, ab, kw. 47. lescol.ti, ab, kw. 48. mevacor.ti, ab, kw. 49. altocor.ti, ab, kw. 50. pravachol.ti, ab, kw. 51. lipostat.ti, ab, kw. 52. zocor.ti, ab, kw. 53. mevinolin.ti, ab, kw. 54. compactin.ti, ab, kw. 55. altoprev.ti, ab, kw. 56. zypitamag.ti, ab, kw. 57. ezallor.ti, ab, kw. |
| **Tolerance** | 58. drug tolerance/ 59. tolerance.ti, ab, kw. 60. intolerance.ti, ab, kw. 61. tolerant.ti, ab, kw. 62. intolerant.ti, ab, kw. 63. tolerability.ti, ab, kw. 64. intolerability.ti, ab, kw. |
| **Combinations** | 65. 1 or 2 or 3 or 4 or 5 or 6 or 7 or 8 or 9 or 10 or 11 66. 12 or 13 or 14 or 15 or 16 or 17 or 18 or 19 or 20 or 21 or 22 or 23 or 24 or 25 or 26 or 27 or 28 or 29 or 30 or 31 or 32 or 33 or 34 or 35 or 36 or 37 67. 38 or 39 or 40 or 41 or 42 or 43 or 44 or 45 or 46 or 47 or 48 or 49 or 50 or 51 or 52 or 53 or 54 or 55 or 56 or 57 68. 58 or 59 or 60 or 61 or 62 or 63 or 64 69. 65 and 66 and 67 and 68 |

**Supplement 3D: Search syntax in CINAHL**

| **Keyword groups** | **Search Syntax in CINAHL** |
| --- | --- |
| **Primary care** | 1. (MH "Primary Health Care") 2. TI (primary care) OR AB (primary care) OR (DE "primary care") 3. TI (primary healthcare or primary health care) OR AB (primary healthcare or primary health care) OR ((DE "Primary Health Care") OR (DE "Primary Healthcare") ) 4. TI (primary medical care) OR AB (primary medical care) OR (DE "Primary Medical Care" ) 5. TI (primary prevention) OR AB (primary prevention) OR (DE "Primary Prevention" ) 6. TI (general practice) OR AB (general practice) OR DE (general practice) 7. TI (general practitioner) OR AB (general practitioner) OR DE (general practitioner) 8. TI (community pharmacy) OR AB (community pharmacy) OR DE (community pharmacy) 9. TI (rural healthcare) OR AB (rural healthcare) OR DE (rural healthcare) 10. TI (district healthcare) OR AB (district healthcare) OR DE (district healthcare) |
| **Disease condition** | 11. (MH "Cardiovascular Disease") OR DE (cardiovascular disease) OR TI (cardiovascular disease) OR AB (cardiovascular disease) 12. (MH "Hyperlipidemia") OR DE (hyperlipidemia) OR TI (hyperlipidemia) OR AB (hyperlipidemia) 13. (MH "Dyslipidemia") OR DE (dyslipidemia) OR TI (dyslipidemia) OR AB (dyslipidemia) 14. DE (hypertension) OR TI (hypertension) OR AB (hypertension) 15. DE (CVD) OR TI (CVD) OR AB (CVD) 16. DE (cardiovascular disease risk factor) OR TI (cardiovascular disease risk factor) OR AB (cardiovascular disease risk factor) 17. DE (angina) OR TI (angina) OR AB (angina) 18. DE (CVD risk factor) OR TI (CVD risk factor) OR AB (CVD risk factor) 19. DE (atherosclerotic cardiovascular disease risk factor) OR TI (atherosclerotic cardiovascular disease risk factor) OR AB (atherosclerotic cardiovascular disease risk factor) 20. DE (hyperlipid) OR TI (hyperlipid) OR AB (hyperlipid) 21. DE (hypercholesterolemia) OR TI (hypercholesterolemia) OR AB (hypercholesterolemia) 22. DE (hypercholesterol) OR TI (hypercholesterol) OR AB (hypercholesterol) 23. DE (familial hyperlipidemia) OR TI (familial hyperlipidemia) OR AB (familial hyperlipidemia) 24. DE (heart disease) OR TI (heart disease) OR AB (heart disease) 25. DE (atherosclerotic disease risk factor) OR TI (atherosclerotic disease risk factor) OR AB (atherosclerotic disease risk factor) 26. DE (cardiovascular risk) OR TI (cardiovascular risk) OR AB (cardiovascular risk) 27. DE (cholesterolemia) OR TI (cholesterolemia) OR AB (cholesterolemia) 28. DE (cardiovascular) OR TI (cardiovascular) OR AB (cardiovascular) 29. DE (cholesterol) OR TI (cholesterol) OR AB (cholesterol) 30. DE (high blood pressure) OR TI (high blood pressure) OR AB (high blood pressure) 31. DE (overweight) OR TI (overweight) OR AB (overweight) 32. DE (obese) OR TI (obese) OR AB (obese) 33. DE (obesity) OR TI (obesity) OR AB (obesity) |
| **Statin** | 34. (MH "Statins") OR DE (statins) OR TI (statins) OR AB (statins) 35. DE (statin) OR TI (statin) OR AB (statin) 36. DE (*statin) OR TI (*statin) OR AB (*statin) 37. DE (hydroxymethylglutaryl-CoA reductase inhibitors) OR TI (hydroxymethylglutaryl-CoA reductase inhibitors) OR AB (hydroxymethylglutaryl-CoA reductase inhibitors) 38. DE (lipid lowering) OR TI (lipid lowering) OR AB (lipid lowering) 39. DE (cholesterol lowering) OR TI (cholesterol lowering) OR AB (cholesterol lowering) 40. DE (lipitor) OR TI (lipitor) OR AB (lipitor) 41. DE (baycol) OR TI (baycol) OR AB (baycol) 42. DE (lescol) OR TI (lescol) OR AB (lescol) 43. DE (mevacor) OR TI (mevacor) OR AB (mevacor) 44. DE (altocor) OR TI (altocor) OR AB (altocor) 45. DE (pravachol) OR TI (pravachol) OR AB (pravachol) 46. DE (lipostat) OR TI (lipostat) OR AB (lipostat) 47. DE (zocor) OR TI (zocor) OR AB (zocor) 48. DE (mevinolin) OR TI (mevinolin) OR AB (mevinolin) 49. DE (compactin) OR TI (compactin) OR AB (compactin) 50. DE (altoprev) OR TI (altoprev) OR AB (altoprev) |
| **Tolerance** | 51. (MH "Drug Tolerance") 52. DE (tolerance) OR TI (tolerance) OR AB (tolerance) 53. DE (intolerance) OR TI (intolerance) OR AB (intolerance) 54. DE (tolerant) OR TI (tolerant) OR AB (tolerant) 55. DE (intolerant) OR TI ( intolerant) OR AB (intolerant) 56. DE (tolerability) OR TI (tolerability) OR AB (tolerability) 57. DE (intolerability) OR TI (intolerability) OR AB (intolerability) |
| **Combinations** | 58. S1 OR S2 OR S3 OR S4 OR S5 OR S6 OR S7 OR S8 OR S9 OR S10 59. S11 OR S12 OR S13 OR S14 OR S15 OR S16 OR S17 OR S18 OR S19 OR S20 OR S21 OR S22 OR S23 OR S24 OR S25 OR S26 OR S27 OR S28 OR S29 OR S30 OR S31 OR S32 OR S33 60. S34 OR S35 OR S36 OR S37 OR S38 OR S39 OR S40 OR S41 OR S42 OR S43 OR S44 OR S45 OR S46 OR S47 OR S48 OR S49 OR S50 61. S51 OR S52 OR S53 OR S54 OR S55 OR S56 OR S57 62. S58 AND S59 AND S60 AND S61 63. S58 AND S59 AND S60 AND S61 |

**Supplement 4: List of excluded studies**

**Studies that did not assess the adverse events or discontinuation of statins due to intolerance (Wrong outcomes):**

1. Cicero, A. F. G., Fogacci, F., Morbini, M., Colletti, A., Bove, M., Veronesi, M., Giovannini, M. and Borghi, C. (2017). Nutraceutical Effects on Glucose and Lipid Metabolism in Patients with Impaired Fasting Glucose: A Pilot, Double-Blind, Placebo-Controlled, Randomized Clinical Trial on a Combined Product. *High Blood Pressure & Cardiovascular Prevention, 24*(3), 283-288. Doi: [10.1007/s40292-017-0206-3](https://dx.doi.org/10.1007/s40292-017-0206-3)
2. Mazza, A., Nicoletti, M., Lenti, S., Torin, G., Rigatelli, G., Pellizzato, M. and Fratter, A. (2021). Effectiveness and Safety of Novel Nutraceutical Formulation Added to Ezetimibe in Statin-Intolerant Hypercholesterolemic Subjects with Moderate-to-High Cardiovascular Risk. *Journal of Medicinal Food, 24*(1), 59-66. Doi: [10.1089/jmf.2020.0019](https://dx.doi.org/10.1089/jmf.2020.0019)
3. Spigoni, V., Aldigeri, R., Antonini, M., Micheli, M. M., Fantuzzi, F., Fratter, A., Pellizzato, M., Derlindati, E., Zavaroni, I., Bonadonna, R. C. and Dei Cas, A. (2017). Effects of a new nutraceutical formulation (Berberine, red yeast rice and chitosan) on non-HDL cholesterol levels in individuals with dyslipidemia: Results from a randomized, double blind, placebo-controlled study. *International Journal of Molecular Sciences, 18*(7). Doi: [10.3390/ijms18071498](https://dx.doi.org/10.3390/ijms18071498)
4. Renner, H. M., Hollar, A., Stolpe, S. F. and Marciniak, M. W. (2017). Pharmacist-to-prescriber intervention to close therapeutic gaps for statin use in patients with diabetes: A randomized controlled trial. Journal of American Pharmacists Association, 57(3s), S236-S242.e1. Doi: [10.1016/j.japh.2017.04.009](https://dx.doi.org/10.1016/j.japh.2017.04.009)
5. Olomu, A., Kelly-Blake, K., Hart-Davidson, W., Gardiner, J., Luo, Z., Heisler, M. and Holmes-Rovner, M. (2022). Improving diabetic patients' adherence to treatment and prevention of cardiovascular disease (Office Guidelines Applied to Practice-IMPACT Study)-a cluster randomized controlled effectiveness trial. *Trials, 23*(1), 659. Doi: [10.1186/s13063-022-06581-6](https://dx.doi.org/10.1186/s13063-022-06581-6)
6. Muckelbauer, Rebecca, Englert, Heike, Rieckmann, Nina, Chen, Chih-Mei, Wegscheider, Karl, Völler, Heinz, Katus, Hugo A, Willich, Stefan N and Müller-Nordhorn, Jacqueline. (2015). Long-term effect of a low-intensity smoking intervention embedded in an adherence program for patients with hypercholesterolemia: Randomized controlled trial. *Preventive Medicine, 77*,155-161. Doi: [10.1016/j.ypmed.2015.05.026](https://dx.doi.org/10.1016/j.ypmed.2015.05.026)
7. Cutler, R.L., Torres-Robles, A., Wiecek, E., Drake, B., Van der Linden, N., Benrimoj, S.I.C. and Garcia-Cardenas, V. (2019). Pharmacist-led medication non-adherence intervention: Reducing the economic burden placed on the Australian health care system. *Patient Preference and Adherence, 13*, 853-862. Doi: [10.2147/PPA.S191482](https://dx.doi.org/10.2147/PPA.S191482)
8. Volpe, M., Pegoraro, V., Peduto, I., Heiman, F. and Meto, S. (2022). Extemporaneous combination therapy with nebivolol/zofenopril in hypertensive patients: usage in Italy. *Current Medical Research and Opinion, 38*(10), 1673-1681. Doi: [10.1080/03007995.2022.2096352](https://dx.doi.org/10.1080/03007995.2022.2096352)
9. Stabile, E., Franzese, M., Chianese, S., Alfani, A., Gerardi, D., Colaiori, I., Annunziata, M., Nappi, P., Scalise, M., Di Serafino, L., Puzone, B., Avvedimento, M., Leone, A., Ilardi, F., Piccolo, R., Franzone, A., Cirillo, P., Morisco, C., Trimarco, B. and Esposito, G. (2021). Predictors of adherence to composite therapy after acute coronary syndromes. *Journal of Cardiovascular Medicine, 22*(8), 645-651. Doi: [10.2459/jcm.0000000000001201](https://dx.doi.org/10.2459/jcm.0000000000001201)
10. Osborn, D., Burton, A., Walters, K., Atkins, L., Barnes, T., Blackburn, R., Craig, T., Gilbert, H., Gray, B., Hardoon, S., Heinkel, S., Holt, R., Hunter, R., Johnston, C., King, M., Leibowitz, J., Marston, L., Michie, S., Morris, R., Morris, S., Nazareth, I., Omar, R., Petersen, I., Peveler, R., Pinfold, V., Stevenson, F. and Zomer, E. (2019). Primary care management of cardiovascular risk for people with severe mental illnesses: the Primrose research programme including cluster RCT. *Programme Grants for Applied Research, 7*(2), vii-98. Doi: [10.3310/pgfar07020](https://dx.doi.org/10.3310/pgfar07020)
11. Muhlestein, J. B., Knowlton, K. U., Le, V. T., Lappe, D. L., May, H. T., Min, D. B., Johnson, K. M., Cripps, S. T., Schwab, L. H., Braun, S. B., Bair, T. L. and Anderson, J. L. (2022). Coronary Artery Calcium Versus Pooled Cohort Equations Score for Primary Prevention Guidance: Randomized Feasibility Trial. *JACC: Cardiovascular Imaging, 15*(5), 843-855
12. Peiris, D., Usherwood, T., Panaretto, K., Harris, M., Hunt, J., Redfern, J., Zwar, N., Colagiuri, S., Hayman, N., Lo, S., Patel, B., Lyford, M., Macmahon, S., Neal, B., Sullivan, D., Cass, A., Jackson, R. and Patel, A. (2015). Effect of a computer-guided, quality improvement program for cardiovascular disease risk management in primary health care: The treatment of cardiovascular risk using electronic decision support cluster-randomized trial. *Circulation: Cardiovascular Quality and Outcomes, 8*(1), 87-95. Doi: [10.1161/CIRCOUTCOMES.114.001235](https://dx.doi.org/10.1161/CIRCOUTCOMES.114.001235)
13. Rinehart, S. N., Collins, C., Glover, J. and Rice, W. M. (2021). Evaluation of a pharmacist-driven medication adherence enhancement service. *Journal of Managed Care and Specialty Pharmacy, 27*(4), 507-515
14. Ferrer Estrela, F., Peris Molina, M.T., Úbeda Pascual, A. and D'Ocon Navaza, M.P. (2015). Algorithm of proceedings in the community pharmacy to optimize the use of statins. *Pharmaceutical Care Espana, 17*(1), 272-286
15. Barnes, B., Hincapie, A. L., Luder, H., Kirby, J., Frede, S. and Heaton, P. C. (2018). Appointment-based models: A comparison of three model designs in a large chain community pharmacy setting. *Journal of American Pharmacists Association, 58*(2), 156-162.e1. Doi: [10.1016/j.japh.2018.01.005](https://dx.doi.org/10.1016/j.japh.2018.01.005)
16. Benson, G. A., Sidebottom, A., Hayes, J., Miedema, M. D., Boucher, J., Vacquier, M., Sillah, A., Gamam, S. and VanWormer, J. J. (2019). Impact of ENHANCED (diEtitiaNs Helping pAtieNts CarE for Diabetes) Telemedicine Randomized Controlled Trial on Diabetes Optimal Care Outcomes in Patients with Type 2 Diabetes. *Journal of the Academy of Nutrition and Dietetics, 119*(4), 585-598. Doi: [10.1016/j.jand.2018.11.013](https://dx.doi.org/10.1016/j.jand.2018.11.013)
17. Bai, J. W., Boulet, G., Halpern, E. M., Lovblom, L. E., Eldelekli, D., Keenan, H. A., Brent, M., Paul, N., Bril, V., Cherney, D. Z., Weisman, A. and Perkins, B. A. (2016). Cardiovascular disease guideline adherence and self-reported statin use in longstanding type 1 diabetes: results from the Canadian study of longevity in diabetes cohort. *Cardiovascular Diabetology, 15*. Doi: [10.1186/s12933-015-0318-9](https://dx.doi.org/10.1186/s12933-015-0318-9)
18. Carter, B. L., Levy, B., Gryzlak, B., Xu, Y., Chrischilles, E., Dawson, J., Vander Weg, M., Christensen, A., James, P. and Polgreen, L. (2018). Cluster-Randomized Trial to Evaluate a Centralized Clinical Pharmacy Service in Private Family Medicine Offices. *Circulation: Cardiovascular Quality & Outcomes, 11*(6), e004188-e004188. Doi: [10.1161/CIRCOUTCOMES.117.004188](https://dx.doi.org/10.1161/CIRCOUTCOMES.117.004188)
19. Barrios, V., Escobar, C., Arrarte, V., García, E., Fernández, M.R., Rincón, L.M. and Roldán, C. (2020). First national registry on the effectiveness and safety of evolocumab in clinical practice in patients attended in cardiology in Spain. The RETOSS-CARDIO study. *Clinica e Investigacion en Arteriosclerosis, 32*(6), 231-241. Doi: [10.1016/j.arteri.2020.05.002](https://dx.doi.org/10.1016/j.arteri.2020.05.002)
20. Gulayin, Pablo E., Lozada, Alfredo, Beratarrechea, Andrea, Gutierrez, Laura, Poggio, Rosana, Chaparro, Raúl Martín, Santero, Marilina, Masson, Walter, Rubinstein, Adolfo and Irazola, Vilma. (2019). An Educational Intervention to Improve Statin Use: Cluster RCT at the Primary Care Level in Argentina. *American Journal of Preventive Medicine, 57*(1), 95-105. Doi: [10.1016/j.amepre.2019.02.018](https://dx.doi.org/10.1016/j.amepre.2019.02.018)
21. Boettiger, D. C., Newall, A. T., Phillips, A., Bendavid, E., Law, M. G., Ryom, L., Reiss, P., Mocroft, A., Bonnet, F., Weber, R., El-Sadr, W., d’Arminio Monforte, A., de Wit, S., Pradier, C., Hatleberg, C. I., Lundgren, J., Sabin, C., Kahn, J. G. and Kazi, D. S. (2021). Cost-effectiveness of statins for primary prevention of atherosclerotic cardiovascular disease among people living with HIV in the United States. *Journal of the International AIDS Society, 24*(3). Doi: 10.1002/jia2.25690
22. Kones, R. (2009). The Jupiter study, CRP screening, and aggressive statin therapy-implications for the primary prevention of cardiovascular disease. *Therapeutic Advances in Cardiovascular Disease, 3*(4), 309-315. Doi: [10.1177/1753944709337056](https://dx.doi.org/10.1177/1753944709337056)
23. Morton, J. I., Liew, D., Watts, G. F., Zoungas, S., Nicholls, S. J., Dixon, P. and Ademi, Z. (2025). Rethinking Cardiovascular Prevention: Cost-Effective Cholesterol Lowering for Statin-Intolerant Patients in Australia and the UK. *European Journal of Preventive Cardiology, 26*
24. Pearson, G. J., Francis, G. A., Romney, J. S., Gilchrist, D. M., Opgenorth, A. and Gyenes, G. T. (2006). The clinical effect and tolerability of ezetimibe in high-risk patients managed in a specialty cardiovascular risk reduction clinic. *Canadian Journal of Cardiology, 22*(11), 939-945. Doi: 10.1016/S0828-282X(06)70313-0
25. Thuraisingham, S., Tan, K. H., Chong, K. S., Yap, S. F. and Pasamanikam, K. (2000). A randomised comparison of simvastatin versus simvastatin and low cholesterol diet in the treatment of hypercholesterolaemia. *International Journal of Clinical Practice, 54*(2), 78-84. Doi: 10.1111/j.1742-1241.2000.tb11854.x
26. Pearson, T. A., Denke, M. A., McBride, P. E., Battisti, W. P., Brady, W. E. and Palmisano, J. (2005). A community-based, randomized trial of ezetimibe added to statin therapy to attain NCEP ATP III goals for LDL cholesterol in hypercholesterolemic patients: the ezetimibe add-on to statin for effectiveness (EASE) trial. *Mayo clinic proceedings, 80*(5), 587-595. Doi: 10.4065/80.5.587
27. Kardas, P. (2013). An education-behavioural intervention improves adherence to statins. *Central European Journal of Medicine, 8*(5), 580-5. Doi: 10.2478/s11536-013-0170-9
28. Stuurman-Bieze, A. G., Hiddink, E. G., Van Boven, J. F. and Vegter, S. (2013). Proactive pharmaceutical care interventions improve patients’ adherence to lipid-lowering medication. *Annals of Pharmacotherapy, 47*(11), 1448-56. Doi: 10.1177/1060028013501146
29. Simpson, S. H., Lin, M. and Eurich, D. T. (2017). Community Pharmacy–Based Inducement Programs Associated With Better Medication Adherence: A Cohort Study. *Annals of Pharmacotherapy, 51*(8), 630-9. Doi: 10.1177/1060028017703720.
30. Párraga-Martínez, I., Escobar-Rabadán, F., Rabanales-Sotos, J., Lago-Deibe, F., Téllez-Lapeira, J. M., Villena-Ferrer, A., Blasco-Valle, M., Ferreras-Amez, J. M., Morena-Rayo, S., Campo, J. M. D. C-D., Ayuso-Raya, M. C and Pérez-Pascual, J. J. (2018). Efficacy of a combined strategy to improve low-density lipoprotein cholesterol control among patients with hypercholesterolemia: a randomized clinical trial. *Revista Española de Cardiología (English Edition), 71*(1), 33-41. Doi: 10.1016/j.rec.2017.05.029
31. Wei, X., Zhang, Z., Chong, M. K., Hicks, J. P., Gong, W., Zou, G., Zhong, J., Walley, J. D., Upshur, R. E. G. and Yu, M. (2021). Evaluation of a package of risk-based pharmaceutical and lifestyle interventions in patients with hypertension and/or diabetes in rural China: A pragmatic cluster randomised controlled trial. *PLoS Medicine, 18*(7), e1003694. Doi: 10.1371/journal.pmed.1003694

**Studies with study designs that did not follow the inclusion criteria of this study (Wrong study design):**

1. Steinhagen-Thiessen, E., Dänschel, W., Buffleben, C., Smolka, W., Pittrow, D. and Hildemann, S.K. (2013). Extended-release niacin/laropiprant for lipid management: Observational study in clinical practice. *International Journal of Clinical Practice, 67*(6), 527-535
2. Taybeh, E. O., Al-Alami, Z. M. and Albasha, A. (2020). Statin use in Jordan: Patients experience and attitude toward adverse drug reactions. *Jordan Journal of Pharmaceutical Sciences, 13*(2), 197-205
3. Minoretti, P., Biagi, M. and Emanuele, E. (2022). An open-label study on the short-term effects of a novel EFSA-compliant nutraceutical combination in mild-to-moderate hypercholesterolemia. *Avicenna Journal of Phytomedicine, 12*(6), 559-565. Doi: [10.22038/ajp.2022.20662](https://dx.doi.org/10.22038/ajp.2022.20662)
4. Shalev, V., Goldshtein, I., Halpern, Y. and Chodick, G. (2014). Association between persistence with statin therapy and reduction in low-density lipoprotein cholesterol level: analysis of real-life data from community settings. *Pharmacotherapy, 34*(1), 1-8. Doi: [10.1002/phar.1326](https://dx.doi.org/10.1002/phar.1326)
5. Jowett, S., Barton, P., Roalfe, A., Fletcher, K., Hobbs, F. D. R., McManus, R. J. and Mant, J. (2017). Cost-effectiveness analysis of use of a polypill versus usual care or best practice for primary prevention in people at high risk of cardiovascular disease. *PLoS ONE, 12*(9)
6. Mefford, M. T., Zhou, M., Zhou, H., Derakhshan, H., Harrison, T. N., Zia, M., Kanter, M. H., Scott, R. D., Imley, T. M., Sanders, M. A., Timmins, R. and Reynolds, K. (2023). Safety Net Program to Improve Statin Initiation Among Adults With High Low-Density Lipoprotein Cholesterol. *American Journal of Preventive Medicine, 65*(4), 687-695. Doi: [10.1016/j.amepre.2023.04.009](https://dx.doi.org/10.1016/j.amepre.2023.04.009)
7. Muhlestein, J. B., Knowlton, K., Le, V. T., Lappe, D., May, H., Min, D., Johnson, K., Cripps, S. T., Schwab, L. H., Braun, S. B., Bair, T. and Anderson, J. L. (2020). Effect on Patient Adherence to Primary Prevention Recommendations for Statin Therapy Based on the National Guidelines-Supported Pooled Cohort Risk Equation or a Coronary Artery Calcium Score: Preliminary Findings from the Vanguard Study for the Corcal Ran. *Journal of the American College of Cardiology, 75*(11)
8. Patterson, J.A., Holdford, D.A. and Saxena, K. (2016). Cost-benefit of appointment-based medication synchronization in community pharmacies. *American Journal of Managed Care, 22*(9), 587-593
9. Arancón-Monge, J. M., de Castro-Cuenca, A., Serrano-Vázquez, Á, Campos-Díaz, L., Rodríguez Barrientos, R. and Del Cura-González, I. (2020). Effects of changing the appearance of medications in safety and adherence in chronic patients over 65 years of age in primary care. CAMBIMED Study. *Atención Primaria, 52*(2), 77-85. Doi: [10.1016/j.aprim.2019.06.001](https://dx.doi.org/10.1016/j.aprim.2019.06.001)
10. Umeda, T., Hayashi, A., Harada, A., Okuyama, K., Baxter, C. A., Tokita, S. and Teramoto, T. (2018). Low-Density Lipoprotein Cholesterol Goal Attainment Rates by Initial Statin Monotherapy Among Patients With Dyslipidemia and High Cardiovascular Risk in Japan - A Retrospective Database Analysis. *Circulation Journal, 82*(6), 1605-1613. Doi: [10.1253/circj.CJ-17-0971](https://dx.doi.org/10.1253/circj.CJ-17-0971)
11. Jia, X., Al Rifai, M., Ramsey, D. J., Ahmed, S. T., Akeroyd, J. M., Nambi, V., Ballantyne, C. M., Petersen, L. A., Stone, N. J. and Virani, S. S. (2019). Association Between Lipid Testing and Statin Adherence in the Veterans Affairs Health System. *The American Journal of Medicine, 132*(9), e693-e700. Doi: [10.1016/j.amjmed.2019.04.002](https://dx.doi.org/10.1016/j.amjmed.2019.04.002)
12. Porath, A., Arbelle, J. E., Fund, N., Cohen, A. and Mosseri, M. (2018). Statin Therapy: Diabetes Mellitus Risk and Cardiovascular Benefit in Primary Prevention. *Israel Medical Association Journal, 20*(8), 480-485
13. Badillo-Alonso, H., Martínez-Alanis, M., Sánchez-Huesca, R., Lerma, A. and Lerma, C. (2023). Effectiveness of the Combination of Enalapril and Nifedipine for the Treatment of Hypertension versus Empirical Treatment in Primary Care Patients. *Journal of Cardiovascular Development and Disease, 10*(6). Doi: 10.3390/jcdd10060243
14. Li, J. H., Joy, S. V., Haga, S. B., Orlando, L. A., Kraus, W. E., Ginsburg, G. S. and Voora, D. (2014). Genetically guided statin therapy on statin perceptions, adherence, and cholesterol lowering: A pilot implementation study in primary care patients. *Journal of Personalized Medicine, 4*(2):147-162. Doi: 10.3390/jpm4020147
15. Albosta, M., Grant, J. K. and Michos, E. D. (2023). Bempedoic Acid: Lipid Lowering for Cardiovascular Disease Prevention. *Heart International, 17*(2), 27-34. Doi: 10.17925/HI.2023.17.2.1
16. Mazhar, F., Hjemdahl, P., Sjölander, A., Kahan, T., Jernberg, T. and Carrero, J. J. (2024). Intensity of and adherence to lipid-lowering therapy as predictors of goal attainment and major adverse cardiovascular events in primary prevention. *American Heart Journal, 269*, 118-130. Doi: 10.1016/j.ahj.2023.12.010
17. Insani, W. N., Whittlesea, C., Ju, C., Man, K., Alwafi, H. H., Alsharif, A., Chapman, S. and Wei, L. (2022). Statin-related adverse drug reactions in UK primary care consultations: A retrospective cohort study to evaluate the risk of cardiovascular events and all-cause mortality. *Pharmacoepidemiology and Drug Safety, 31*(Supplement 2), 293-294
18. Khan, S., Holbrook, A. and Shah, B. R. (2018). Does Googling lead to statin intolerance? International Journal of Cardiology, 262, 25-27. Doi: 10.1016/j.ijcard.2018.02.085
19. Descamps, O.S. (2013). Mona lisa study: Effect of a preparation of red yeast rice (Artchol®) in a cohort of hypercholesterolemic patients treated by general practitioners. *Louvain Medical, 132*(4), 149-154
20. Eriksson, M., Hådell, K., Holme, I., Walldius, G. and Kjellström, T. (1998). Compliance with and efficacy of treatment with pravastatin and cholestyramine: a randomized study on lipid-lowering in primary care. *Journal of Internal Medicine,243*(5), 373-80. Doi: 10.1046/j.1365-2796.1998.00294.x
21. McCormack, T., Harvey, P., Gaunt, R., Allgar, V., Chipperfield, R. and Robinson, P. (2010). Incremental cholesterol reduction with ezetimibe/simvastatin, atorvastatin and rosuvastatin in UK General Practice (IN-PRACTICE): Randomised controlled trial of achievement of Joint British Societies (JBS-2) cholesterol targets. *International Journal of Clinical Practice, 64*(8), 1052-1061. Doi: 10.1111/j.1742-1241.2010.02429.x
22. Raal, F., Schamroth, C., Patel, J. and Becker, P. (2007). A multicentre, open-label, observational local study to evaluate the low-density lipoprotein cholesterol-lowering effect of ezetimibe as prescribed in daily routine practice in the South African population. *Cardiovascular Journal of Africa, 18*(5), 325-9
23. Wald, D. S. and Wald, N. J. (2012). Implementation of a simple age-based strategy in the prevention of cardiovascular disease: The Polypill approach. Journal of Evaluation in Clinical Practice, 18(3):612-615. Doi: 10.1111/j.1365-2753.2011.01637.x

**Studies that included patients who were prescribed statins for secondary prevention of CVD (Wrong population):**

1. Cho, L. (2023). Bempedoic Acid and Cardiovascular Outcomes in Statin Intolerant Patients At High Cardiovascular Risk: CLEAR OUTCOME. *Journal of Clinical Lipidology, 17*(4), e62-e62. Doi: [10.1016/j.jacl.2023.05.090](https://dx.doi.org/10.1016/j.jacl.2023.05.090)
2. Ansbro, E., Masri, S., Prieto-Merino, D., Willis, R., Aoun Bahous, S., Molfino, L., Boulle, P. and Perel, P. Fixed dose combination drugs for cardiovascular disease in a prolonged humanitarian crisis in Lebanon: An implementation study. *BMJ Open, 13*(1)
3. Volpp, K. G., Troxel, A. B., Mehta, S. J., Norton, L., Zhu, J., Lim, R., Wang, W., Marcus, N., Terwiesch, C., Caldarella, K., Levin, T., Relish, M., Negin, N., Smith-McLallen, A., Snyder, R., Spettell, C. M., Drachman, B., Kolansky, D. and Asch, D. A. (2017). Effect of Electronic Reminders, Financial Incentives, and Social Support on Outcomes After Myocardial Infarction: The HeartStrong Randomized Clinical Trial. *JAMA Internal Medicine, 177*(8), 1093-1101. Doi: [10.1001/jamainternmed.2017.2449](https://dx.doi.org/10.1001/jamainternmed.2017.2449)
4. Sotorra-Figuerola, G., Ouchi, D., García-Sangenís, A., Giner-Soriano, M. and Morros, R. (2022). Pharmacological treatment after acute coronary syndrome: Baseline clinical characteristics and gender differences in a population-based cohort study. *Atención Primaria, 54*(1), 102157. Doi: [10.1016/j.aprim.2021.102157](https://dx.doi.org/10.1016/j.aprim.2021.102157)
5. Anderson, J. L., Knowlton, K. U., May, H. T., Bair, T. L., Armstrong, S. O., Lappé, D. L. and Muhlestein, J. B. (2018). Temporal changes in statin prescription and intensity at discharge and impact on outcomes in patients with newly diagnosed atherosclerotic cardiovascular disease-Real-world experience within a large integrated health care system: The IMPRES study. *Journal of Clinical Lipidology, 12*(4), 1008-1018.e1. Doi: [10.1016/j.jacl.2018.03.084](https://dx.doi.org/10.1016/j.jacl.2018.03.084)
6. Booth, J. N., Colantonio, L. D., Chen, L., Rosenson, R. S., Monda, K. L., Safford, M. M., Kilgore, M. L., Brown, T. M., Taylor, B., Dent, R., Muntner, P. and Levitan, E. B. (2017). Statin Discontinuation, Reinitiation, and Persistence Patterns Among Medicare Beneficiaries After Myocardial Infarction: A Cohort Study. *Circulation: Cardiovascular Quality and Outcomes, 10*(10). Doi: [10.1161/circoutcomes.117.003626](https://dx.doi.org/10.1161/circoutcomes.117.003626)
7. Iqbal, S., Sabbour, H. M., Ashraf, T., Santos, R. D. and Buckley, A. (2024). First Report of Inclisiran Utilization for Hypercholesterolemia Treatment in Real-world Clinical Settings in a Middle East Population. *Clinical Therapeutics, 46*(3), 186-93.
8. Yamane, S. S., De Gagne, J. C., Riggs, A., Kimberly, G. D. and Holye, M. (2020). Assessment of a patient-centered initiative to improve hypertension management for adults with comorbid type 2 diabetes at a free clinic in the rural south. *Nursing Forum, 55*(3), 348-355. Doi: [10.1111/nuf.12434](https://dx.doi.org/10.1111/nuf.12434)
9. Di Martino, M., Alagna, M., Lallo, A., Gilmore, K. J., Francesconi, P., Profili, F., Scondotto, S., Fantaci, G., Trifirò, G., Isgrò, V., Davoli, M. and Fusco, D. (2021). Chronic polytherapy after myocardial infarction: the trade-off between hospital and community-based providers in determining adherence to medication. *BMC Cardiovascular Disorders, 21*(1). Doi: [10.1186/s12872-021-01969-9](https://dx.doi.org/10.1186/s12872-021-01969-9)
10. Olomu, A., Hart-Davidson, W., Luo, Z., Kelly-Blake, K. and Holmes-Rovner, M. (2016). Implementing shared decision making in federally qualified health centers, a quasi-experimental design study: The Office-Guidelines Applied to Practice (Office-GAP) program. *BMC Health Services Research, 16*(1). Doi: [10.1186/s12913-016-1603-3](https://dx.doi.org/10.1186/s12913-016-1603-3)
11. Kassner, U., Hollstein, T., Grenkowitz, T., Scharnagl, H., Maerz, W. and Steinhagen-Thiessen, E. (2018). PCSK9-Inhibitor treatment of cardiovascular high risk patients in a real-world setting. *European Heart Journal, 39*, 1041
12. Di Martino, M., Alagna, M., Cappai, G., Mataloni, F., Lallo, A., Perucci, C. A., Davoli, M. and Fusco, D. (2016). Adherence to evidence-based drug therapies after myocardial infarction: is geographic variation related to hospital of discharge or primary care providers? A cross-classified multilevel design. *BMJ Open, 6*(4), e010926.Doi: [10.1136/bmjopen-2015-010926](https://dx.doi.org/10.1136/bmjopen-2015-010926)
13. Heng, W. K., Ng, Y. P., Ooi, G. S., Hat, H. B., Jamaluddin, N. B., Nawi, N. A. B. M., Hasim, H. B. and Wahab, N. B. (2019). Comparison of the efficacy and level of adherence for morning versus evening versus before bedtime administration of simvastatin in hypercholesterolemic patients. *Medical Journal of Malaysia, 74*(6), 477-482
14. Berthold, H. K., Unverdorben, S., Degenhardt, R., Bulitta, M. and Gouni-Berthold, I. (2006). Effect of policosanol on lipid levels among patients with hypercholesterolemia or combined hyperlipidemia: A randomized controlled trial. *JAMA, 295*(19), 2262-2269. Doi: 10.1001/jama.295.19.2262
15. Den Hartog, F. R., Van Kalmthout, P. M., Van Loenhout, T. T., Schaafsma, H. J., Rila, H. and Verbeugt, F. (2001). Pravastatin in acute ischaemic syndromes: Results of a randomised placebo-controlled trial. *International Journal of Clinical Practice, 55*(5), 300-304. Doi: 10.1111/j.1742-1241.2001.tb11043.x
16. Enajat, M., Teerenstra, S., Van Kuilenburg, J. T., Van Sorge-Greve, A. H. N., Albers-Akkers, M. T. H., Verheugt, F. W. A. and Pop, G. A. M. (2009). Safety of the combination of intensive cholesterol-lowering therapy with oral anticoagulation medication in elderly patients with atrial fibrillation: A randomized, double-blind, placebo-controlled study. *Drugs and Aging, 26*(7), 585-593. Doi: 10.2165/10558450-000000000-00000

**Studies with interventions that did not align with the inclusion criteria of this study (Wrong intervention):**

**Studies which comparator groups that did not align with the inclusion criteria of this study (Wrong comparator):**

1. Roshandel, G., Khoshnia, M., Poustchi, H., Hemming, K., Kamangar, F., Gharavi, A., Ostovaneh, M.R., Nateghi, A., Majed, M., Navabakhsh, B., Merat, S., Pourshams, A., Nalini, M., Malekzadeh, F., Sadeghi, M., Mohammadifard, N., Sarrafzadegan, N., Naemi-Tabiei, M., Fazel, A., Brennan, P., Etemadi, A., Boffetta, P., Thomas, N., Marshall, T., Cheng, K.K. and Malekzadeh, R. (2019). Effectiveness of polypill for primary and secondary prevention of cardiovascular diseases (PolyIran): a pragmatic, cluster-randomised trial. *The Lancet, 394*(10199), 672-683. Doi: 10.1016/S0140-6736(19)31791-X
2. Avellone, G., Di Garbo, V., Guarnotta, V., Scaglione, R., Parrinello, G., Purpura, L., Torres, D. and Campisi, D. (2010). Efficacy and safety of long-term ezetimibe/simvastatin treatment in patients with familial hypercholesterolemia. *International Angiology, 29*(6), 514-524
3. Fras, Z. and Mikhailidis, D. P. (2008). Statin plus ezetimibe treatment in clinical practice: The SI-SPECT (Slovenia (SI) Statin Plus Ezetimibe in Cholesterol Treatment) monitoring of clinical practice study. *Current Medical Research and Opinion, 24*(9), 2467-2476. Doi: 10.1185/03007990802303772

**Supplement 5:** **Risk of bias and quality assessment of included studies**

| **Study** | **D1** | **D2** | **D3** | **D4** | **D5** | **Overall** |
| --- | --- | --- | --- | --- | --- | --- |
| Thongtang et al. (2020) |  |  |  |  |  |  |
| Nissen et al. (2023) |  |  |  |  |  |  |
| Laufs et al. (2020) |  |  |  |  |  |  |
| Kennedy et al. (2011) |  |  |  |  |  |  |
| Malekzadeh et al. (2010) |  |  |  |  |  |  |

**Supplement 5A:** **Quality assessment of randomized controlled trials using the RoB 2 tool**

D1: Randomization process; D2: Deviations from intended interventions; D3: Missing outcome data; D4: Measurement of outcome; D5: Selection of reported results.

High risk	Some concern	Low risk

Study quality of RCTs (as percentage)

|  | Low risk | Some concerns | High risk |
| --- | --- | --- | --- |
| Randomization process | 80 | 20 | 0 |
| Deviation from intended interventions | 80 | 20 | 0 |
| Missing outcome data | 100 | 0 | 0 |
| Measurement of outcome | 100 | 0 | 0 |
| Selection of reported results | 20 | 80 | 0 |
| Overall | 20 | 80 | 0 |

**Supplement 5B: Quality assessment of nonrandomized studies of interventions using the ROBINS-I tool**

| **Study** | **D1** | **D2** | **D3** | **D4** | **D5** | **D6** | **D7** | **Overall bias** |
| --- | --- | --- | --- | --- | --- | --- | --- | --- |
| Solomon et al. (2020) |  |  |  |  |  |  |  |  |
| Hong et al. (2022) |  |  |  |  |  |  |  |  |
| Iqbal et al (2022) |  |  |  |  |  |  |  |  |
| Meek et al. (2012) |  |  |  |  |  |  |  |  |

D1: Confounding; D2: Selection of participants into the study; D3: Classification of interventions; D4: Deviations from intended interventions; D5: Missing outcome data; D6: Measurement of outcome; D7: Selection of reported results.

High risk	Some concern	Low risk

Study quality of NRSIs (as percentage)

|  | Low risk | Some concerns | High risk |
| --- | --- | --- | --- |
| Confounding | 0 | 100 | 0 |
| Selection of participants into the study | 100 | 0 | 0 |
| Classification of interventions | 0 | 50 | 50 |
| Deviations from intended interventions | 100 | 0 | 0 |
| Missing data | 100 | 0 | 0 |
| Measurement of outcomes | 0 | 100 | 0 |
| Selection of the reported result | 50 | 50 | 0 |
| Overall bias | 0 | 50 | 50 |

**Supplement 6: Subgroup analysis (forest plots of RCTs) for statin intolerance**

**Supplement 6A: Role of participant age in the effectiveness of intervention**

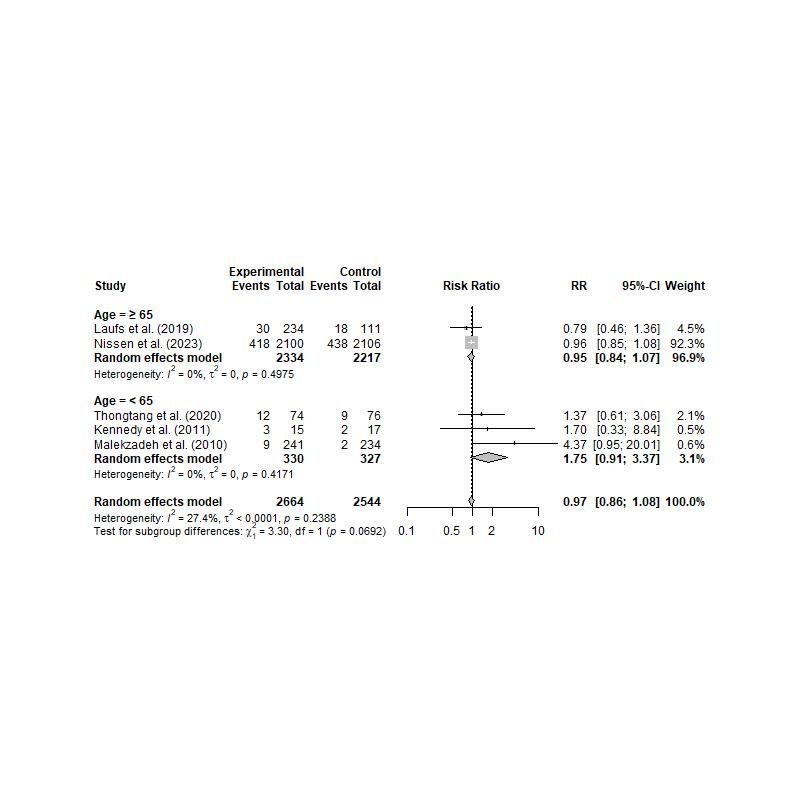

**Supplement 6B: Role of geographic location in the effectiveness of intervention**

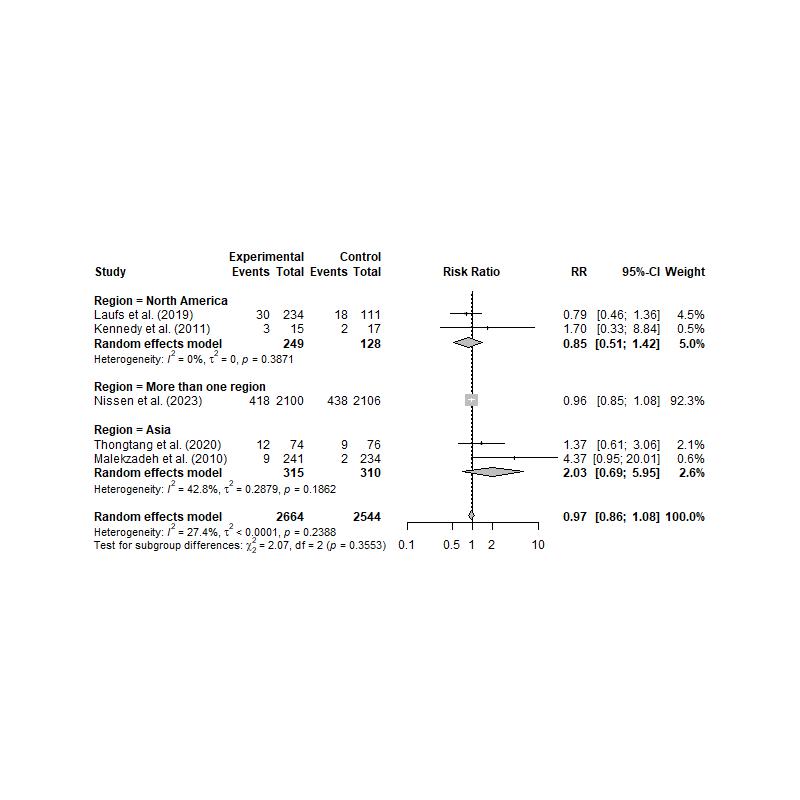

**Supplement 7: Subgroup analysis (forest plots of all studies including RCTs and NRSIs) for statin intolerance**

**Supplement 7A: Role of intervention strategy in the effectiveness of intervention**

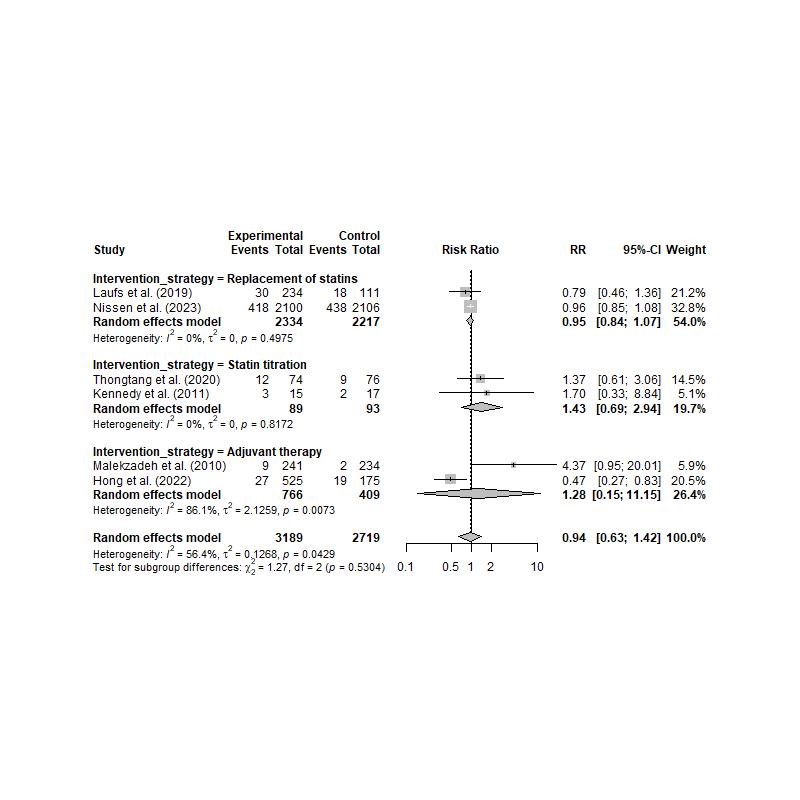

**Supplement 7B: Role of participant age in the effectiveness of intervention**

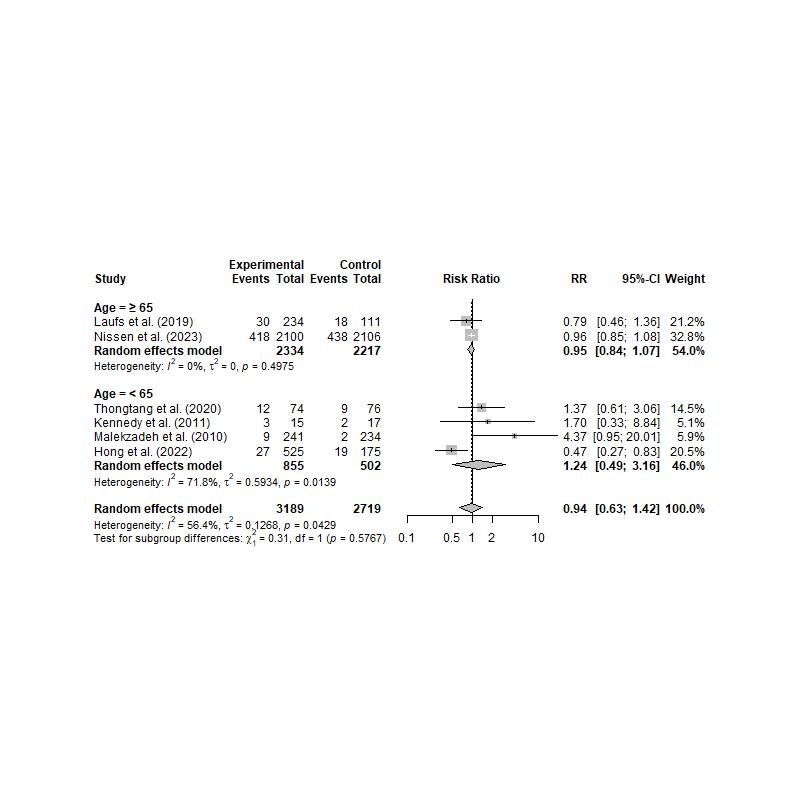

**Supplement 7C: Role of geographic location in the effectiveness of intervention**

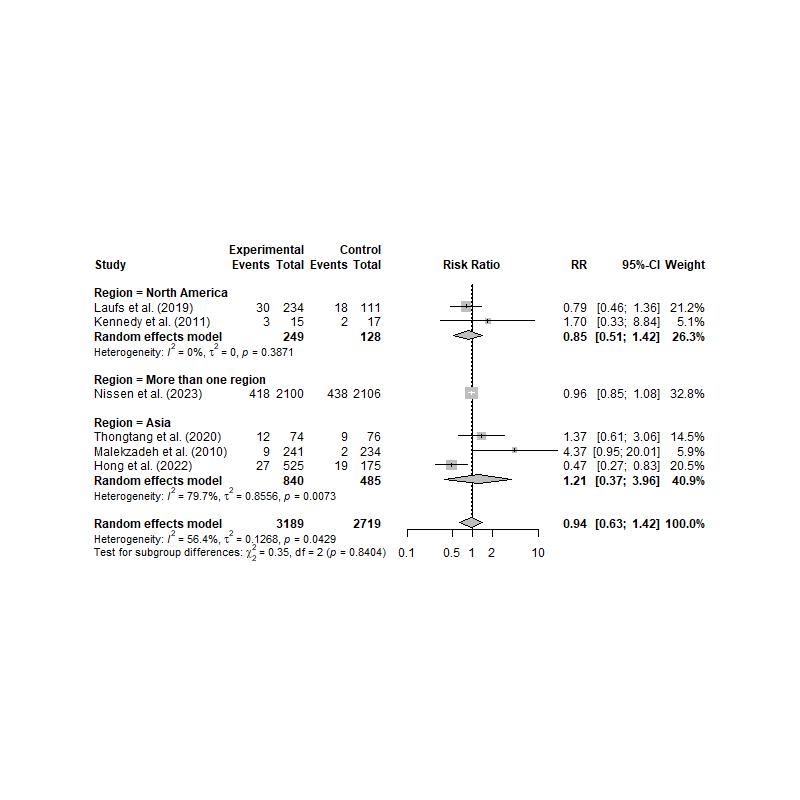

**Supplement 8: Details of all outcomes**

| **Study** | **Intervention** | | **Control** | |
| --- | --- | --- | --- | --- |
|  | **N** | **n (%)** | **N** | **n (%)** |
| ***Adverse events*** | | | | |
| Thongtang et al. (2020) | 74 | 12 (16.22) | 76 | 9 (11.84) |
| Nissen et al. (2023) | 2,100 | 1,785 (85.00) | 2,106 | 1,744 (82.81) |
| Laufs et al. (2020) | 234 | 51 (21.79) | 111 | 20 (18.02) |
| Hong et al. (2022) | 525 | 27 (5.14) | 175 | 19 (10.86) |
| ***Serious adverse events*** | | | | |
| Nissen et al. (2023) | 2,100 | 418 (19.90) | 2,106 | 438 (20.80) |
| Laufs et al. (2020) | 234 | 14 (5.98) | 111 | 4 (3.60) |
| ***Adverse drug reactions*** | | | | |
| Laufs et al. (2020) | 234 | 51 (21.79) | 111 | 20 (18.02) |
| Hong et al. (2022) | 525 | 11 (2.09) | 175 | 0 (0.00) |
| ***Myalgia*** | | | | |
| Nissen et al. (2023) | 2,100 | 88 (4.19) | 2,106 | 124 (5.89) |
| Laufs et al. (2020) | 234 | 11 (4.70) | 111 | 8 (7.21) |
| Kennedy et al. (2011) | 15 | 3 (20.00) | 17 | 2 (11.76) |
| ***Muscle symptoms*** | | | | |
| Thongtang et al. (2020) | 74 | 12 (16.22) | 76 | 9 (11.84) |
| Laufs et al. (2020) | 234 | 30 (12.82) | 111 | 18 (16.22) |
| Hong et al. (2022) | 525 | 8 (1.52) | 175 | 4 (2.89) |
| ***Creatine kinase > 10 ULN*** | | | | |
| Nissen et al. (2023) | 2,100 | 5 (0.24) | 2,106 | 2 (0.09) |
| Malekzadeh et al. (2010) | 241 | 9 (3.73) | 234 | 2 (0.85) |

**Supplement 9: Subgroup analysis (forest plots) for statin discontinuation**

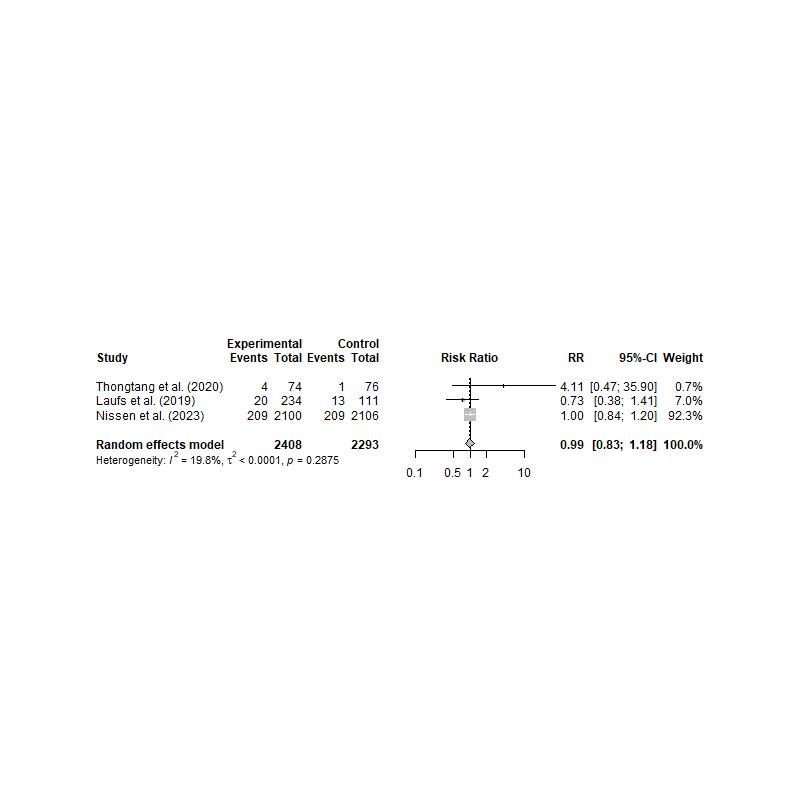
